# Population-level indirect effects of HPV vaccination in high-income settings: a Bayesian modelling study

**DOI:** 10.64898/2026.11.13.26367966

**Authors:** Runze Wang, Mark Jit

**Affiliations:** Department of Biostatistics, School of Global Public Health, New York University, New York, NY, United States of America; Department of Global and Environmental Health, School of Global Public Health, New York University, New York, NY, United States of America

**Keywords:** HPV vaccination, Indirect protection, Herd immunity, Bayesian hierarchical modelling

## Abstract

**Background:** Human papillomavirus (HPV) vaccination provides direct protection to vaccinated individuals, but also indirectly protects unvaccinated individuals through reduced HPV transmission. Mechanistic transmission models that predict the magnitude of such effects have been crucial to informing vaccination strategies. However, nearly two decades of post-vaccination experience in high-income countries now allow us to empirically characterise how population-level indirect effects vary with vaccine coverage, time since vaccine introduction, and age.

**Methods:** We synthesised studies reporting HPV16/18 prevalence before and after vaccine introduction in high-income settings between 2007 and 2025. Indirect effects were estimated as the observed reduction in HPV16/18 prevalence after accounting for expected direct vaccine protection. We used a Bayesian hierarchical model to examine associations with vaccine coverage, time since vaccine introduction, and age at effect assessment across routine, catch-up, and non-targeted vaccination groups. XGBoost was used as a complementary, non-parametric approach.

**Findings:** Data from 11 high-income settings were included. Indirect effects increased during the first five to eight years after vaccine introduction and then remained relatively stable. Indirect effects generally decreased with increasing vaccine coverage. Patterns with age differed across vaccination groups. In the routine group, indirect effects increased during adolescence and remained positive at older observed ages. In the catch-up group, indirect effects decreased with age and approached zero or became negative after approximately age 25 years. XGBoost identified similar non-linear patterns.

**Interpretation:** Indirect protection against HPV16/18 develops gradually after vaccine introduction and varies with vaccine coverage, age, and vaccination group. This empirical approach can complement transmission models by using real-world evidence to characterise population-level indirect protection. However, making predictions outside the populations and ranges represented in the data should be done cautiously.

**Funding:** This study received no specific funding.

**Research in context:** *Evidence before this study:* We searched PubMed for studies published up to Oct 26, 2026. The search terms were (“HPV vaccin” [All Fields] OR “human papillomavirus vaccin” [All Fields]) AND (“herd immunity” [All Fields] OR “herd effect” [All Fields] OR “indirect effect” [All Fields] OR “indirect protection” [All Fields] OR “herd protection” [All Fields]). We included systematic reviews, meta-analyses, observational studies, and modelling studies that assessed the indirect effects of HPV vaccination on HPV infection or HPV-related outcomes. We excluded studies that focused exclusively on direct vaccine protection or economic evaluations that included herd immunity without separately estimating or reporting indirect effects. We identified 301 articles, of which 77 met the inclusion criteria: 52 observational studies, 22 modelling studies, and three systematic reviews or meta-analyses. From those studies, two systematic reviews by Drolet and colleagues (2015 and 2019) synthesised evidence from 14 high-income countries and provided evidence of herd protection following vaccine introduction. A meta-analysis of results from 16 transmission dynamic models by Brisson and colleagues (2016) projected substantial herd effects under different vaccination coverage levels. More recently, an observational study by Meglic and colleagues (2026) used nationwide registry data from Sweden and found age-varying herd effects, with lower incidence of high-grade cervical lesions among unvaccinated women in cohorts exposed to school-based vaccination. Despite growing real-world evidence from high-income countries over nearly two decades, few studies have examined how indirect protection varies with vaccination coverage, time since programme introduction, and age across settings.

*Added value of this study:* To our knowledge, this is the first study to synthesise real-world HPV16/18 prevalence data from 11 high-income settings to estimate the indirect effects of HPV vaccination over the period 2007–2025. Our findings show that indirect protection generally increases during the first several years after vaccine introduction before stabilising, with different patterns across age groups and vaccination programmes.

*Implications of all the available evidence:* As countries work towards cervical cancer elimination, understanding the indirect protection provided by HPV vaccination is important for assessing its broader population-level impact. This multi-country evidence complements transmission-dynamic models and provides an empirical basis for evaluating the long-term effects of HPV vaccination programmes.

## Introduction

Human papillomavirus (HPV) is the most common sexually transmitted infection ^1^ Persistent infection with high-risk HPV types is a necessary cause of cervical cancer and can also cause other anogenital and oropharyngeal cancers, together accounting for approximately 630,000 new cancer cases worldwide each year.^2^ Since the first HPV vaccine was licensed in 2006, national vaccination programmes have been introduced in many countries and have substantially reduced vaccine-type HPV infection and HPV-related disease.^3^ HPV vaccines with progressively broader type coverage have since become available, including bivalent vaccines targeting HPV16/18,^4^ quadrivalent vaccines additionally targeting HPV6/11,^5^ and nonavalent vaccines protecting against five additional oncogenic HPV types.^6^

High-income countries (HICs) such as Australia, the United Kingdom, and the United States were among the first to implement national HPV vaccination programmes, achieving high vaccine coverage.^7–9^ Most programmes have adopted school-based routine vaccination targeting adolescent girls, with some countries additionally introducing catch-up vaccination for older cohorts who were not eligible for routine immunisation.^8,9^ More recently, many countries have extended these programmes to gender-neutral vaccination.^10–12^ Large clinical trials have established the high efficacy of HPV vaccines in preventing persistent vaccine-type HPV infection and related precancerous lesions among vaccinated individuals.^5,6,13,14^

Beyond direct protection among vaccinated individuals, HPV vaccination can also reduce HPV transmission and consequently lower infection risk among unvaccinated populations through indirect (or herd, or community) protection. In this study, we use the term indirect effect to describe the reduction in HPV infection attributable to reduced transmission beyond the expected direct protection conferred by vaccination.

Substantial population-level reductions in vaccine-type HPV infection and genital warts have been observed in both vaccine-eligible and unvaccinated age groups following the introduction of HPV vaccination programmes.^9,11,15,16^ However, such observational studies have been conducted in only a small number of countries, most of them high-income. Moreover, evidence on the impact of HPV vaccination is most valuable prospectively, for anticipating the benefits that a planned programme is likely to deliver. Findings from other countries may not be generalisable, owing to differences in HPV epidemiology, population demography, sexual behaviour, interventions such as screening, and prevalence of co-morbidities such as HIV.

To address these limitations, transmission-dynamic models have been widely used to project the long-term direct and indirect effects of HPV vaccination under different vaccination scenarios, particularly in settings where sufficient post-vaccination evidence from real-world vaccination programmes is not yet available.^17^ However, these models require detailed parameters describing HPV transmission, such as sexual partner change rates and rates governing disease natural history, which can be difficult to measure and may vary across populations.^17^

Nearly two decades after the introduction of HPV vaccination, substantial observational data have accumulated, allowing us to statistically identify patterns in long-term vaccine impact, particularly in high-income countries where the most data are available. The direct effect of vaccination is relatively straightforward to calculate, since individuals naïve to HPV infection are almost completely protected from infection with vaccine-type HPV.^18^ The indirect effect is more complex but can be related to readily measurable population characteristics, such as vaccine coverage, age, and time since vaccine introduction using statistical models.

We therefore synthesised population-level evidence from HICs during 2007–2025 to quantify and characterise the indirect effects of HPV vaccination on HPV prevalence. We used two complementary analytical approaches. The primary analysis used a Bayesian hierarchical model relating indirect effects with key explanatory variables (time since vaccine introduction, age at effect assessment, vaccine coverage, and vaccination-eligibility group), while accounting for heterogeneity across countries and studies. We additionally applied XGBoost as a flexible machine-learning approach to explore whether it identified qualitatively similar patterns without imposing prespecified associations between these factors and indirect effects. The model results can indicate whether there is an association between indirect effects and the explanatory variables explored that is consistent enough to use to predict vaccine effects in settings prior to vaccine introduction.

## Methods

### Literature review

Two systematic reviews provide comprehensive summaries of population-level post-vaccine introduction studies of the impact of vaccination on HPV infections.^11,19^ They contain data from 14 high-income countries with approximately 60 million individuals. The studies included in the two systematic reviews were published between 2007 and 2018.

To extend the existing evidence to later years, we conducted a rapid PubMed search of population-level studies of HPV vaccine impact on HPV prevalence in high-income countries published between October 11, 2018, and July 20, 2025, using the same search strategy as the original reviews (Appendix Part 1).

### Data collection

We extracted data from each study in the review on the population-level impact of the HPV vaccine. Other outcomes examined in the reviews (anogenital warts and high-grade cervical lesions) were not included as they were downstream outcomes of prior HPV infection and had longer latency periods before diagnosis. By the time of this study, six highly efficacious prophylactic HPV vaccines have been licensed worldwide, including three bivalent, two quadrivalent, and one nonavalent vaccine.^4–6,14,20,21^ All the licensed HPV vaccines had two HPV types in common that they protected against: HPV 16 and 18,^22^ the two most common types in cervical cancer. Accordingly, analyses were restricted to HPV16/18 prevalence. To ensure comparability between the pre- and post-vaccination periods, we included only studies that reported HPV prevalence for both periods using the same recruitment methods and population sources. The final inclusion criteria and search results are detailed in Appendix Part 1. Our final dataset included studies from 11 high-income regions, covering millions of individuals between 2007 and 2025.^23–50^

We manually extracted data on pre- and post-vaccination HPV prevalence, corresponding 95% confidence intervals (where available), vaccine coverage, age groups, study periods, and sample sizes for both periods as reported from tables, the main text, or the supplementary materials of the original studies. For studies that presented data in figures without corresponding numerical values, we extracted the corresponding values using the WebPlotDigitizer.^51^ Data extraction was conducted in a Microsoft Excel 2021 workbook. Since studies reported age-specific HPV prevalence by age intervals with lengths that differed between studies, we disaggregated the broader age intervals into single-year age intervals to ensure consistency in analysis. We applied an exponentially damped linear model fitted to study data using Markov chain Monte Carlo (MCMC) methods to interpolate the age-specific post-vaccination prevalence for HPV types 16 and 18.^52^ This approach provided point estimates and 95% credible intervals for each single-year age group (Appendix Part 2.2). In one study that reported only prevalence ratios (PRs), we modified the data provided by Hermida et al. as described in Appendix Part 2.1 to estimate the corresponding prevalence.^53^

Study populations were classified into three groups based on their age and the national vaccination programme in each country (Appendix Part 8): vaccinees in ages eligible for routine vaccination group, vaccinees in ages eligible for catch-up vaccination, vaccinees in ages not targeted by the national immunisation programme (but where some individuals may still be vaccinated, e.g. in the private sector). For age ranges that included populations from more than one of the three groups, the group with the most individuals from the age group was assigned to that age group. When articles reported age-specific vaccine coverage, these data were extracted directly from the studies. If age-specific coverage data were not available from the articles, we obtained vaccine coverage from national immunisation programme reports or publicly available government health statistics, and subsequently reconstructed vaccination-eligibility group-specific coverage following the method described in Appendix Part 2.3. To account for uncertainty in age-specific HPV prevalence estimates obtained from the fitted model, we propagated the uncertainty to our indirect effect estimates using a Monte Carlo approach (Appendix Part 3).

At the population level, we defined the estimated indirect effect of HPV vaccination at age a as the difference between the total effect and the expected direct effect. Here, P_pre_(a) and P_post_(a) indicate HPV16/18 prevalence at age a during the pre- and post-vaccination periods, respectively. C(a) denotes vaccine coverage among individuals aged a at the post-vaccination assessment. The total effect T(a) was defined as the observed absolute reduction in HPV16/18 prevalence for individuals at age a: T(a) = P_pre_(a) − P_post_(a).

The expected direct effect D(a) at age a was estimated from pre-vaccination HPV16/18 prevalence at that age, vaccine coverage at that age, and modified intention-to-treat vaccine effectiveness V_mITT_(a) at the same age: D(a) = P_pre_(a) × C(a) × V_mITT_(a). The estimated indirect effect I(a) was therefore: I (a) = P_pre_(a) − P_post_(a) − P_pre_(a) × C(a) × V_mITT_(a).

Thus, the estimated indirect effect in a particular age group represents the component of the observed population-level reduction in HPV16/18 prevalence in that age group that remained after subtracting the expected direct protection attributable to vaccination (calculated using vaccine coverage in women who have reached that age). Modified intention-to-treat vaccine effectiveness at age a was calculated from intention-to-treat vaccine effectiveness and post-vaccination HPV16/18 prevalence at the same age: V_mITT_(a) = Vm_ITT_ × [1 − P_post_(a)]. Substituting this expression into the indirect-effect equation gives: I(a) = P_pre_(a) − P_post_(a) − P_pre_(a) × C(a) × V_mITT_ × [1 − P_post_(a)]. Prevalence and vaccine coverage were expressed as proportions when calculating indirect effects, which represent absolute differences in prevalence. Negative values indicate that the observed reduction in HPV16/18 prevalence was smaller than the expected direct effect under the assumptions used to construct the indirect-effect estimate. Indirect effect estimates and their standard errors were expressed in percentage points for modelling and reporting. We assumed V_mITT_ = 1 (100%), based on previous evidence.^54^

### Modelling

We used a Bayesian hierarchical model to characterise the associations of the estimated indirect effect with vaccine coverage, time since vaccine introduction, and age at effect assessment. Let Y_i_ denote the estimated indirect effect for observation i, with corresponding standard error SE_i_. We specified the following Student’s t-distribution for the likelihood: 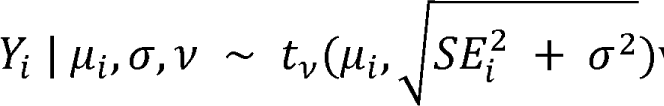 where μ_i_ is the expected indirect effect, σ represents additional residual variation beyond the known uncertainty in each indirect-effect estimate, and ν is the degrees-of-freedom parameter.

Let j_i_, c_i_, and z_i_ indicate the vaccination-eligibility group, country, and study associated with observation i, respectively. The expected indirect effect was modelled as:

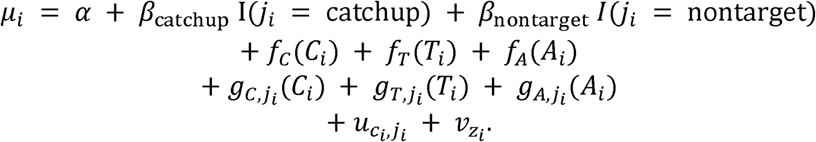

where C_i_, T_i_, and A_i_ represent vaccine coverage, time since vaccine introduction, and age at effect assessment, respectively. The functions f_C_(·), f_T_(·), and f_A_(·) represent the overall smooth associations, whereas g_C,j_(·), g_T,j_(·), and g_A,j_(·) are group-specific smooth terms that allow the shapes of these associations to differ across the three vaccination-eligibility groups. I(.) is an indicator function. All smooth terms were represented using thin-plate regression splines. The terms u_c□,j□_ and v_z□_ are country-by-vaccination-eligibility-group and study-level random intercepts, respectively.

The model was fitted using the Hamiltonian Monte Carlo (HMC) algorithm with four chains, 4,000 iterations, 1,000 warm-up iterations, and a maximum tree depth of 13 in Stan via the brms package. Convergence was assessed visually using trace plots (Appendix Part 5.2). All analyses were conducted in R version 4.4.3. Further details of the model can be found in Appendix Part 5.1.

We also applied XGBoost regression as a complementary analysis to examine the associations between the estimated indirect effect and vaccine coverage, time since vaccine introduction, and age at effect assessment. XGBoost was used because it can flexibly capture complex non-linear associations without specifying their functional forms. It also outperforms other machine learning methods for small datasets.^55^ Given the relatively small sample size in this study, we implemented two model-evaluation approaches to the XGBoost to reduce the risk of overfitting. First, we applied LOOCV (Leave-One-Out Cross-Validation), where each time the model is trained on (n-1) samples and leaves out one sample for testing, repeated n times until every sample has been used for testing. Second, the dataset was split into 70% for training and 30% for testing. An exhaustive grid search was used to identify the optimal hyperparameters for the XGBoost with the highest R-squared value. We generated a SHapley Additive exPlanations (SHAP) dependence plot to quantify the contribution of each predictor to the prediction of the indirect effect.^56^ The analysis was conducted using R version 4.4.3.

### Sensitivity analysis

We examined the study-level random intercepts from the Bayesian hierarchical model to assess residual heterogeneity between studies. Additionally, to evaluate the robustness of the results to the inclusion of countries with larger sample sizes, we performed sensitivity analyses under two scenarios: a model using data from the US and the UK only, and a model excluding data from both those countries.

## Results

Eligible studies identified from the previous systematic reviews and our updated search were conducted in 11 high-income national or subnational settings: the United States, the United Kingdom, Scotland, Denmark, Sweden, Norway, Australia, Japan, Finland, Spain, and the Netherlands (Fig. 1A–B). The type of HPV vaccine used varied across countries and over time, with several countries transitioning from bivalent or quadrivalent to nonavalent vaccines (see details in Appendix Part 8).

**Fig. 1.**
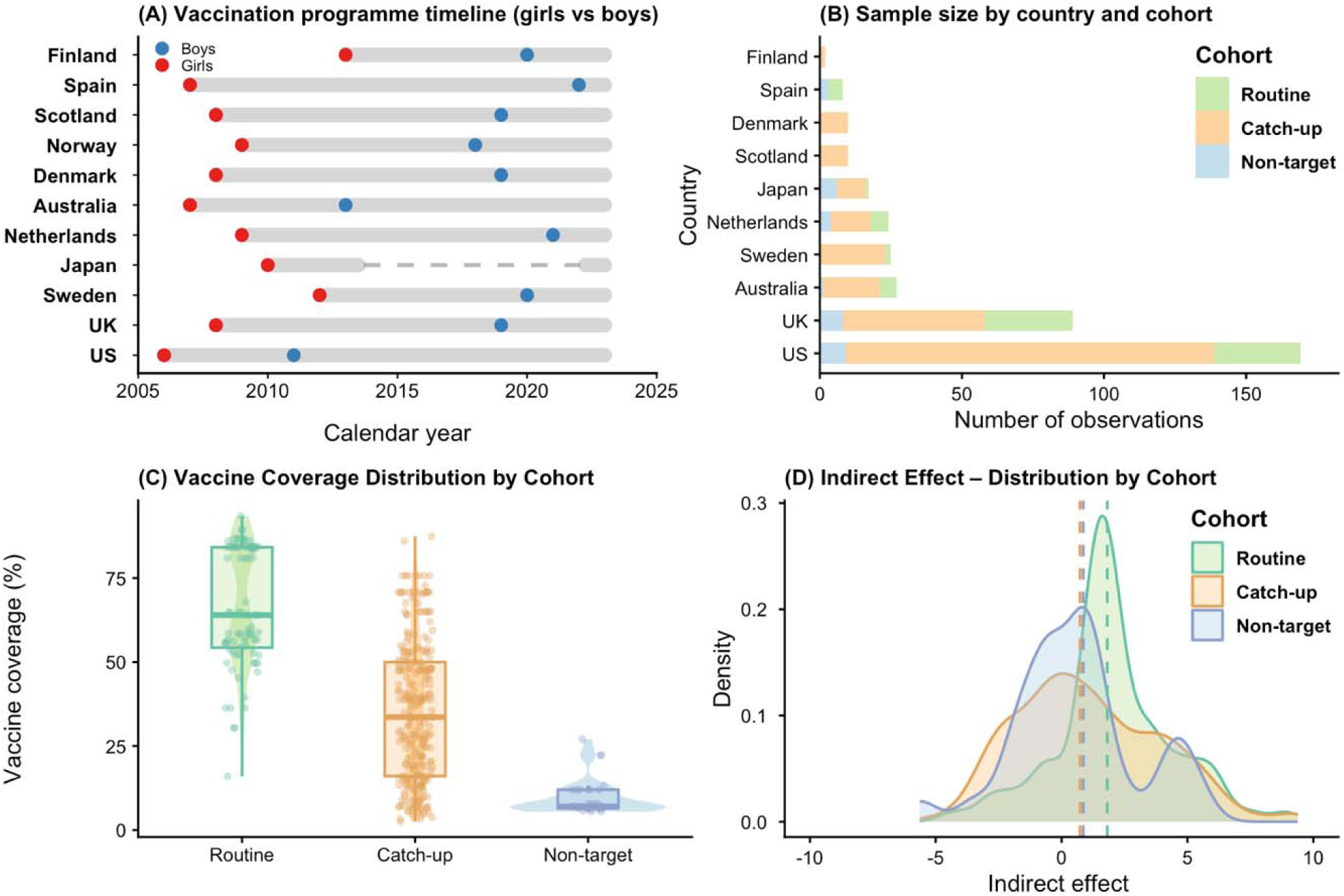
Overview of the data included in the modelling analyses. (A) Calendar year of HPV vaccination programme introduction for girls (red dots) and boys (blue dots) and subsequent use (thick gray lines) across included countries. The dashed line for Japan indicates the temporary suspension of the national HPV programme. (B) Number of data points included in the modelling analyses, stratified by country and vaccination-eligibility group. (C) Distribution of vaccine coverage (%) across routine, catch-up, and non-targeted groups. (D) Distribution of the estimated indirect effect across routine, catch-up, and non-targeted groups. Colours represent vaccination group: routine (green), catch-up (orange), and non-targeted (blue).

In the overall sample, 70·6% of observations were from the catch-up vaccination group, 8·1% from the non-targeted group, and 21·3% from the routine vaccination group. The country distribution was highly unbalanced, with the United States and the United Kingdom together accounting for more than half of the sample. The mean vaccine coverage across all groups was 30·7% (SD = 26·8) (Figure 1C). The routine vaccination group showed the highest median vaccine coverage of 64% (IQR: 54·3–84·2), followed by the catch-up group (33·6%, IQR: 16·1–50), and non-targeted group (7%, IQR: 6·5–12). The mean time since vaccine introduction was 6·29 years (SD = 2·82) (Appendix Part 4).

The Bayesian hierarchical model estimated non-linear associations between indirect effects and vaccine coverage, time since vaccine introduction, and age at effect assessment (Figure 2). Posterior distributions for the model coefficients can be found in Appendix Part 5.3. At the overall population level, estimated indirect effects decreased with increasing vaccine coverage. In contrast, indirect effects increased over the first six to eight years following vaccine introduction before subsequently plateauing. Indirect effects decreased with increasing age, became negative after approximately 25 years, reached a minimum at around 30 years, and subsequently increased towards zero at older ages.

**Fig. 2.**
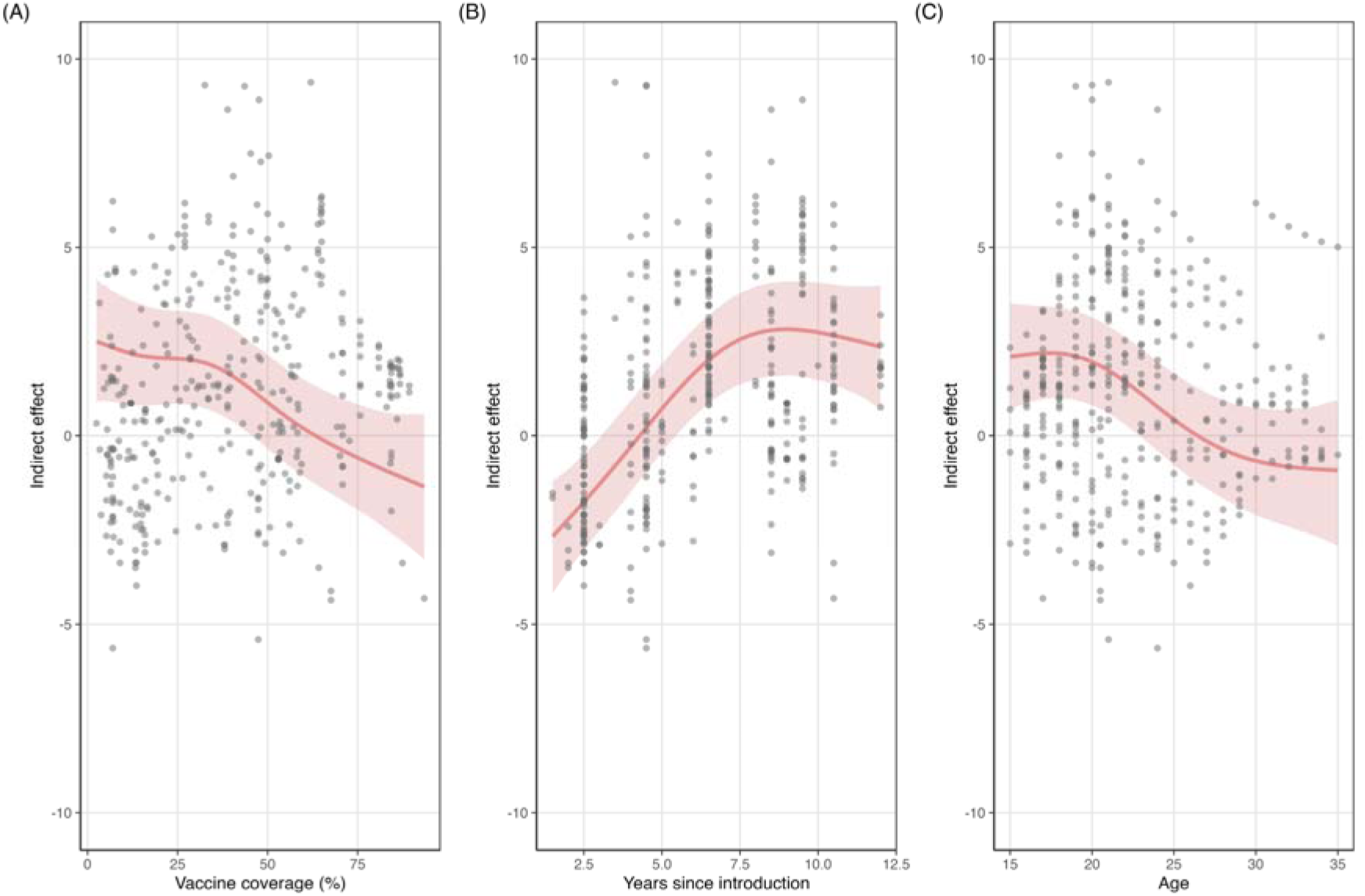
Associations between estimated indirect effects and (A) vaccine coverage, (B) time since vaccine introduction, and (C) age at effect assessment. Solid red curves represent posterior mean estimates from the fitted Bayesian hierarchical model, and shaded ribbons represent 95% credible intervals. Grey points represent the observed study-level estimates.

The associations between indirect effects and the three predictors differed across vaccination-eligibility groups (Figure 3). In all three groups, indirect effects increased with time since vaccine introduction for the first five to six years and subsequently remained stable. In the routine and catch-up vaccinated populations, indirect effects declined with increasing vaccine coverage. In the routine group, indirect effects decreased from strongly positive values at lower coverage levels to approximately zero at higher coverage levels. A similar but weaker declining pattern was observed in the catch-up group. In the non-targeted population, indirect effects also declined with increasing vaccine coverage. However, credible intervals were wider at higher coverage levels because of the limited number of observations. In the routine group, estimated indirect effects increased with age at effect assessment during adolescence, peaked at approximately 18–22 years, and remained positive and relatively stable at older ages. In the catch-up group, indirect effects decreased with increasing age at effect assessment, from positive values at younger ages to approximately zero or negative values after age 25 years. In the non-targeted group, indirect effects increased from negative values at younger ages towards zero at approximately 20–25 years and became positive and relatively stable at older ages.

**Fig. 3.**
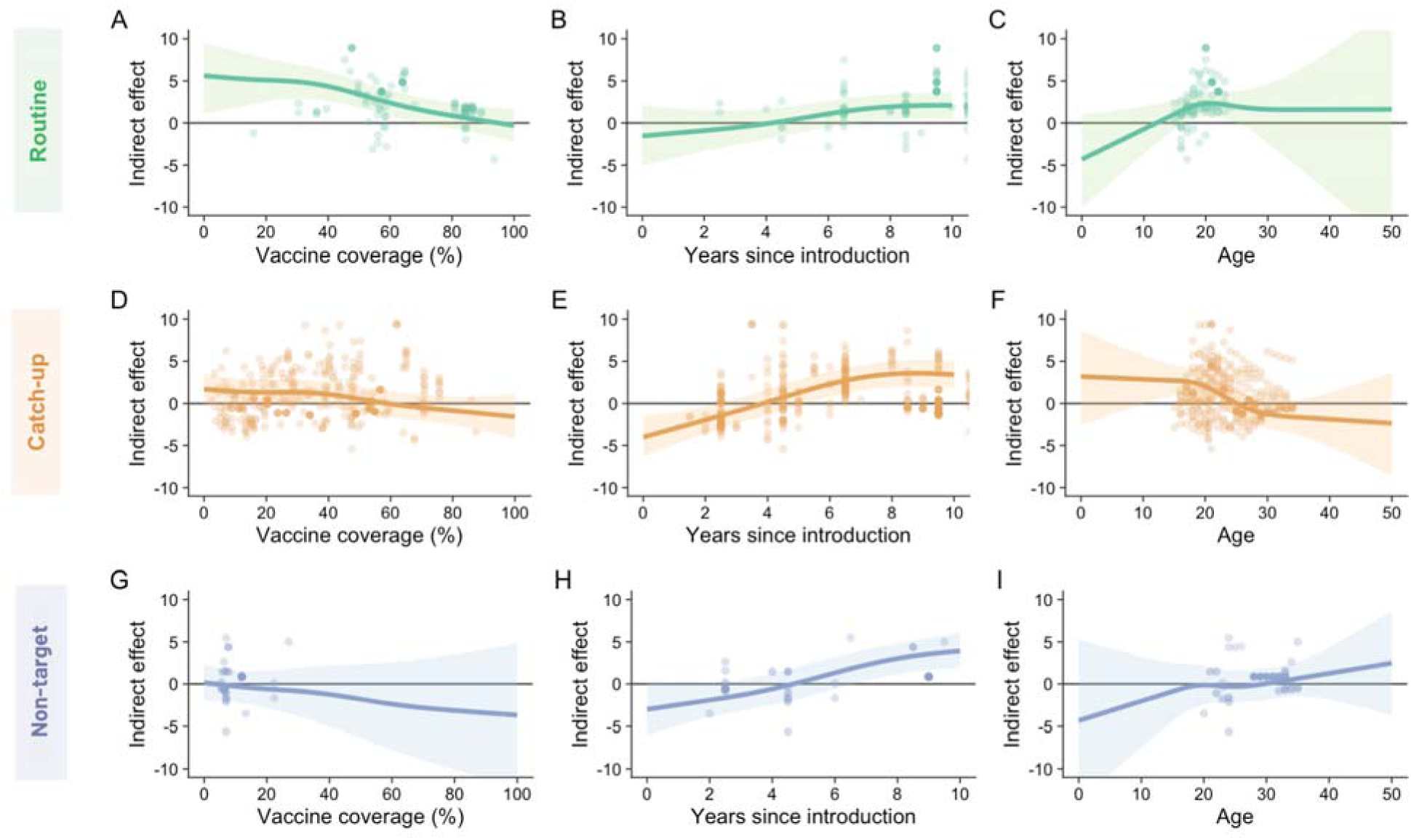
Associations between estimated indirect effects and vaccine coverage, time since vaccine introduction, and age at effect assessment, stratified by vaccination-eligibility group. Solid curves represent posterior mean model estimates and shaded ribbons represent 95% credible intervals. Points represent the indirect-effect estimates included in the analysis. Point opacity reflects the standard error of each estimate, with more opaque points indicating smaller standard errors.

We applied the fitted Bayesian model to a hypothetical programme with 50% vaccine coverage, routine vaccination at age 12, and catch-up vaccination through age 18 (Table 1). In the routine vaccination group, predicted indirect effects generally increased across older ages and later assessment points, changing from negative estimates at younger ages and earlier years following vaccine introduction to positive estimates at older ages and later years. The catch-up group showed a similar pattern.

**Table 1.** Model-predicted indirect effects under a hypothetical HPV vaccination programme with 50% vaccine coverage, routine vaccination at age 12, and catch-up vaccination through age 18.

| Vaccination group | Age at effect assessment | Years since vaccine introduction | Predicted indirect effect (95% CrI) |
|---|---|---|---|
| Routine | 15 | 4 | –0·209 (–2·231 to 1·821) |
| Routine | 18 | 6 | 2·152 (0·332 to 4·000) |
| Routine | 20 | 8 | 3·369 (1·608 to 5·217) |
| Routine | 22 | 10 | 3·499 (1·698 to 5·411) |
| Catch-up | 18 | 4 | 0·926 (–0·507 to 2·356) |
| Catch-up | 20 | 6 | 2·600 (1·175 to 4·023) |
| Catch-up | 22 | 8 | 2·993 (1·536 to 4·421) |
| Catch-up | 24 | 10 | 2·082 (0·576 to 3·567) |

Predictions assume 50% vaccine coverage. Values are population-level posterior mean predictions from the Bayesian hierarchical model with 95% credible intervals (CrI). Indirect effects are expressed in percentage points.

Country-level heterogeneity was modelled through country-specific random intercepts within the hierarchical model. This allowed partial pooling of information across countries while permitting country-specific deviations from the overall population-level association. We further explored group-specific associations between indirect effects and vaccine coverage, time since vaccine introduction, and age across the represented countries.

As shown in Figure 4, country-specific estimates varied in magnitude across vaccination-eligibility groups. In the routine group, Australia, the UK, and the US generally showed the highest estimated indirect effects, whereas Japan and Spain showed the lowest estimates. Similar cross-country variation was observed in the catch-up group, with Australia, Denmark, and the UK showing consistently positive indirect effects. In the non-targeted group, Australia and the UK generally showed higher indirect effects, whereas Japan and Spain showed lower estimates. Because the model included country-by-vaccination-eligibility-group random intercepts but not country-specific smooth functions, countries within each eligibility group shared the same shape for the estimated association between age and indirect effects.

**Fig. 4.**
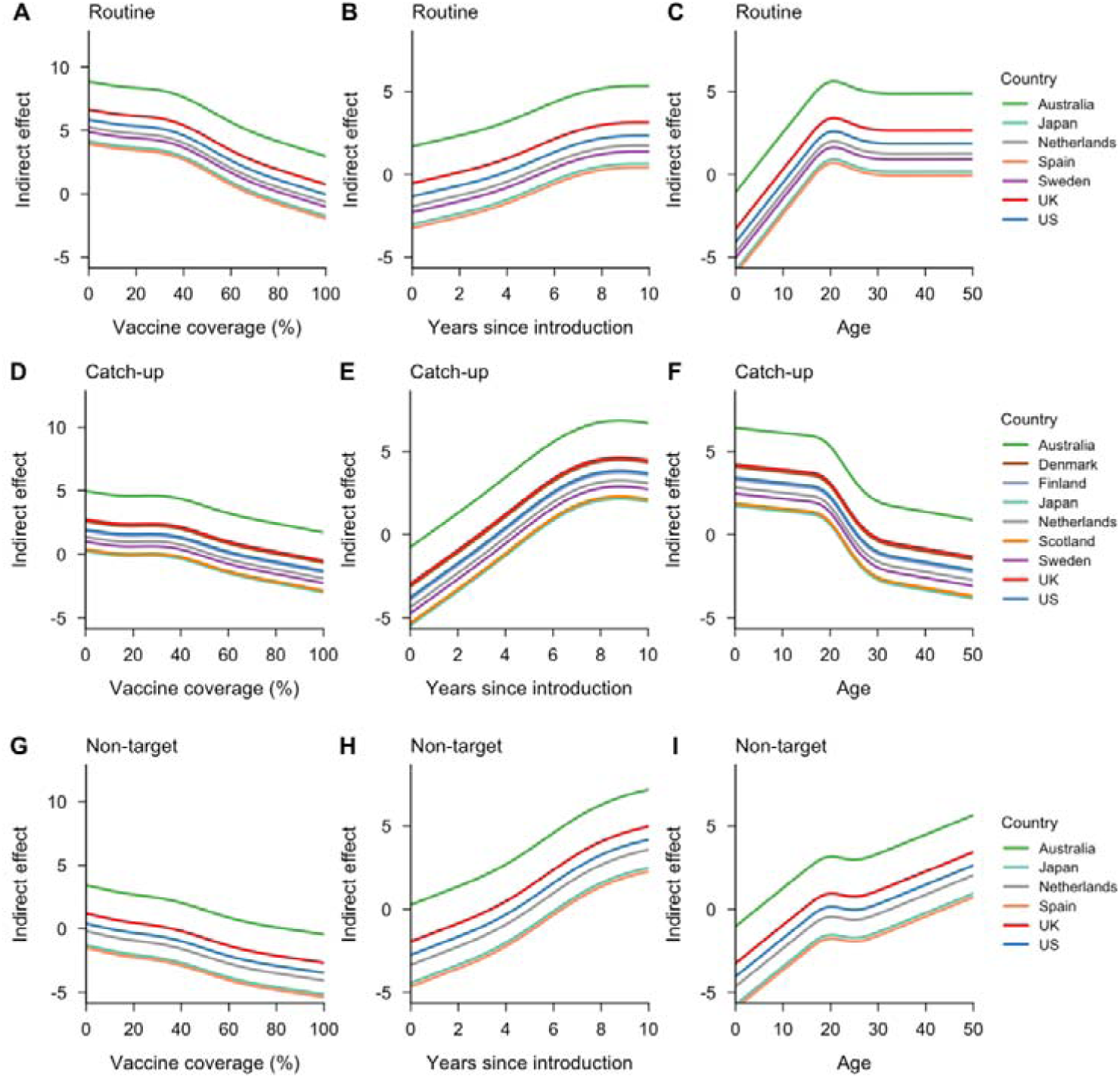
Country-specific partial pooling estimates of indirect vaccine effects from the model. Panels show country-specific posterior mean predictions of indirect vaccine effects across vaccine coverage, years since vaccine introduction, and age, respectively, obtained from the Bayesian hierarchical model with partial pooling across country-by-vaccination-eligibility groups.

We examined the study-level random intercepts from the hierarchical model to assess heterogeneity across studies (Appendix Part 7). Several studies showed positive or negative deviations from the overall mean, indicating that unexplained study-level variability is present even after controlling for covariates and country-level heterogeneity.

### XGBoost results

The XGBoost analysis identified non-linear patterns that resembled those observed in the Bayesian model (Figure 5). SHAP values indicate how much each predictor contributes to the model prediction relative to the average prediction. Larger mean absolute SHAP values indicate greater predictor importance in the model. Years since vaccine introduction had the largest mean absolute SHAP value (1·229), followed by vaccine coverage (0·778) and age at effect assessment (0·723), indicating that time since vaccine introduction had the greatest average contribution to the XGBoost predictions. Vaccine coverage showed a non-linear pattern, with SHAP values varying at lower and intermediate coverage levels and generally declining at higher levels of vaccine coverage. SHAP values for years since vaccine introduction increased rapidly during approximately the first six to seven years and then remained relatively stable. Age at effect assessment showed a more complex non-linear pattern. SHAP values increased until approximately 19–20 years of age, then declined to their lowest level at around 29–30 years and increased thereafter.

**Fig. 5.**
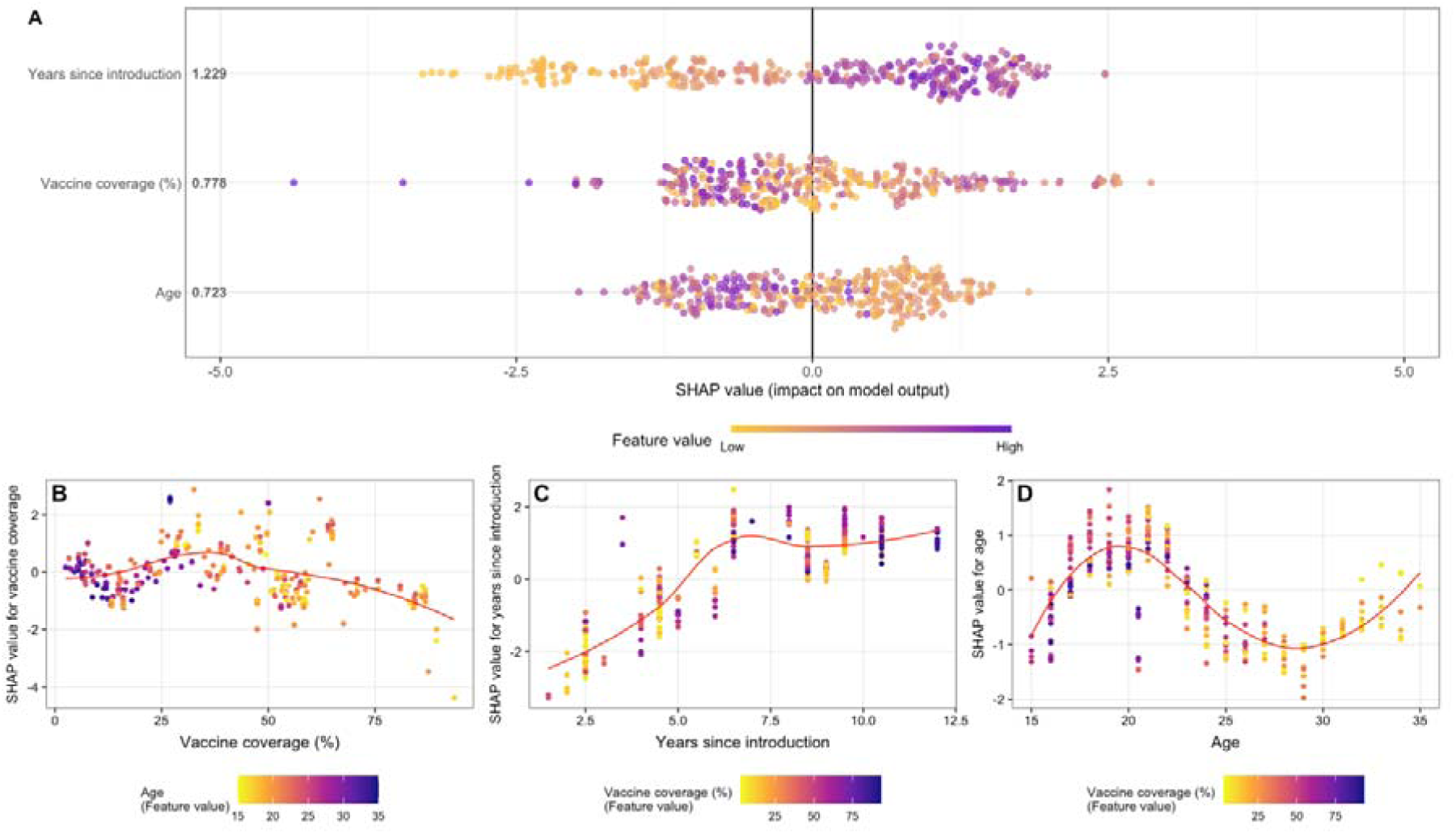
SHAP summary and dependence plots from the complementary XGBoost analysis. (A) SHAP summary plot showing the distributions of SHAP values for years since vaccine introduction, vaccine coverage, and age at effect assessment. Features are ranked according to their mean absolute SHAP values. Each point represents the SHAP value of one predictor for one observation. Positive SHAP values indicate that the observed predictor value shifted the XGBoost-modelled indirect effect above the model’s expected prediction. Negative values indicate that it shifted the modelled indirect effect below the expected prediction. Point colours represent feature values from low to high. SHAP dependence plots are shown for (B) vaccine coverage, (C) years since vaccine introduction, and (D) age at effect assessment. In panels B–D, point colours represent the values of the interacting features indicated by the corresponding colour bars. Red lines represent LOESS (locally estimated scatterplot smoothing) curves fitted to the SHAP values to summarise the overall patterns.

### Sensitivity analysis

In the sensitivity analyses, the overall patterns of associations between estimated indirect effects and vaccine coverage, time since vaccine introduction, and age at effect assessment were broadly similar to those observed in the primary analysis. Some differences were observed in the shape and uncertainty of the associations when data from the US and the UK were either excluded or analysed separately (Appendix Part 6). Both sensitivity analyses suggested that the primary model results were robust to the influence of countries contributing larger numbers of observations.

## Discussion

Nearly two decades after the first HPV vaccine was licensed, HPV vaccination programmes have expanded globally and have substantially reduced vaccine-type HPV infection and related disease. High-income countries were among the earliest adopters and have now accumulated more than a decade of population-level observational evidence on the impact of HPV vaccination.^57,58^ However, despite increasing evidence of herd protection following HPV vaccination, less is known about how indirect protection varies across age, vaccine coverage, and time since programme introduction in real-world settings.

In this study, we synthesised population-level evidence on the indirect effects of HPV vaccination using observational data from multiple high-income settings between 2007 and 2025. We used statistical and machine-learning approaches to characterise patterns of indirect protection across countries and vaccination-eligibility groups. Our findings showed that indirect effects are not static but develop gradually following vaccination programme implementation. Estimated indirect effects increased during the early years following vaccine introduction and plateaued after approximately six to eight years. Second, indirect effects had non-monotonic patterns with age at effect assessment and differed across vaccination-eligibility groups. Third, estimated indirect effects generally decreased as vaccination coverage increased. The XGBoost analysis identified qualitatively similar patterns and provided complementary exploratory evidence.

These associations are consistent with epidemiological reasoning. In our analysis, the indirect effect was defined as the reduction in HPV16/18 prevalence that remained after accounting for expected direct protection among vaccinated individuals (assuming 100% vaccine effectiveness in HPV-naïve individuals). With increasing vaccine coverage, a larger proportion of the observed reduction is assigned to direct vaccine protection, leaving less of a reduction to be attributed to indirect effects. As more time elapses since vaccination, an increasing number of birth cohorts are vaccinated, increasing indirect effects. The observed association between indirect effects and age is less straightforward to interpret because the age range was restricted within some vaccination-eligibility groups. For example, all 81 observations in the routine-vaccination group were assessed at ages 15–24 years. Estimates outside the observed age range for a given group therefore rely on model extrapolation and should be interpreted cautiously.

These relationships mostly concur with results from previous empirical and modelling studies. While indirect effects from vaccination have been observed in older non-vaccinated groups of women, the observed effects have been small, and only appear many years after vaccination and in countries with high vaccine coverage.^11^ The association between indirect effects and coverage has been explored by modelling studies. One such study found that increasing vaccine coverage among 12-year-old girls reduces the impact of indirect effects, although this was limited to routine vaccination groups.^59^ Another modelling study predicted that indirect effects peak at intermediate coverage levels and subsequently decline.^60^

Age-specific patterns of estimated indirect effects differed across vaccination-eligibility groups. In the routine-vaccination group, indirect effects increased during adolescence, and subsequently remained relatively stable at older ages, possibly reflecting increasing levels of sexual activity in older adolescents. In contrast, in the catch-up group, indirect effects decreased with increasing age at effect assessment and approached zero or became negative after approximately 25 years of age. Differences in vaccine coverage and time since vaccination may also contribute to heterogeneity across settings. Findings for the non-targeted population were less clearly interpretable. Although these age groups were not eligible for vaccination through national immunisation programmes, some individuals may still have received vaccination through private insurance or out-of-pocket payment. Vaccination opportunities, exposure histories, and age distributions also varied within this group. In addition, relatively few observations were available at some predictor values. Regions with few or no data points often showed a negative association between coverage and indirect effects. This is epidemiologically implausible and is likely an artefact of extrapolating smooth functions beyond the limits of data. Therefore, findings for the non-targeted population and outside parameter ranges with data points should be interpreted with caution.

Both the Bayesian hierarchical model and XGBoost identified similar associations between indirect effects, vaccine coverage, age at effect assessment, and time since vaccine introduction. The Bayesian model explicitly accounted for the uncertainty of the study-level indirect-effect estimates and enabled partial pooling across settings, while providing interpretable estimates of the associations. In contrast, XGBoost provided a flexible approach for identifying non-linear associations without prespecifying their functional forms. However, its results are less directly interpretable.

A major strength of this study is that we synthesise real-world population-level evidence across high-income countries from 2007 to 2025. Although most primary studies reported HPV prevalence using broad age intervals, we used a model-based approach based on typical age-dependent HPV prevalence patterns to obtain more refined age-specific prevalence estimates.^52^ This allowed us to distinguish indirect effects across routine, catch-up, and non-targeted populations and to identify differences that would not be apparent from aggregated age groups. Additionally, the XGBoost analysis identified similar patterns with the Bayesian method, providing complementary evidence across different modelling approaches.

There are several limitations to this study. First, indirect effects cannot be directly observed and so were estimated as the residual reduction in HPV16/18 prevalence after accounting for expected direct vaccine protection. The estimate may therefore be confounded by time-dependent differences between the pre- and post-vaccination populations. Second, we interpolate age-specific prevalence due to coarseness in reported age-dependent data; the reconstructed age-specific prevalence may not fully reflect the true age distributions. Third, vaccine coverage was not measured in the same way across all studies and settings. Some estimates were reported directly, whereas others were reconstructed from national programme statistics. For example, in some US data, vaccination status in NHANES was self-reported and may therefore be affected by recall bias.^61^ When reconstructing age-specific vaccine coverage in several settings, we used coverage at the final year of programme eligibility and assumed that it remained constant as the cohort aged. In practice, coverage may change slightly over time due to private sector vaccination at later ages. Similarly, some individuals classified as non-targeted may have received vaccination outside national immunisation programmes. Fourth, the included studies differed in their sampling frames, recruitment methods, and laboratory assays. Some were based on subnational populations and may not represent the corresponding national populations.^37,46^ In several studies, long intervals separated the pre- and post-vaccination periods. Changes in HPV testing technologies or diagnostic sensitivity during these periods may have introduced additional measurement heterogeneity.^62^ Although study- and setting-level random effects accounted for some heterogeneity, they could not eliminate systematic differences in study design and measurement. Fifth, our model did not include behavioural covariates such as partner-change rates, age-dependent partnership formation, and age-specific sexual behaviour due to lack of comparable national data.^11,63^ These factors may contribute to differences in indirect protection across countries and populations and may partly explain residual heterogeneity between settings. Finally, our findings were based on population-level observational data collected between 2007 and 2025 from 11 high-income national or subnational settings. The estimated relationships therefore reflect the vaccination programmes, demographic and behavioural characteristics, and healthcare systems of these populations. We need to be cautious to extrapolate to low- and middle-income countries (LMICs) as these factors may differ substantially. Moreover, social and sexual behaviours and healthcare delivery may change over time, even within the same country. Caution is also needed when extrapolating beyond the observed ranges of vaccine coverage, time since vaccine introduction, and age at effect assessment.

Our empirical approach complements mechanistic HPV transmission models. Dynamic transmission models can simulate HPV transmission under different vaccination scenarios and project population-level effects beyond observed follow-up periods. However, they require behavioural and epidemiological parameters that can be difficult to measure consistently across settings.^17^ In contrast, our approach uses observed post-vaccination data to relate estimated indirect effects to measurable population-level characteristics, including vaccine coverage, age at effect assessment, and time since vaccine introduction. The residual country-level and study-level heterogeneity we found suggests that the model may be unreliable for extrapolating to settings without post-vaccination HPV prevalence data. Hence our approach cannot completely replace mechanistic models that simulate transmission processes and project effects beyond the observed data. Nevertheless, it can still provide a broad indication of the likely magnitude of HPV vaccine impact, particularly when data, time, or expertise required for mechanistic modelling are unavailable.

Future studies could extend this approach to more HPV types, such as the additional oncogenic types covered by the nonavalent vaccine, and to clinical outcomes, such as anogenital warts and high-grade cervical lesions. Such analyses would face additional challenges. Nonavalent vaccines have seen a shorter history of use, so there would be fewer data points with which to fit models. Vaccine impact on clinical outcomes would take longer to observe than impact on HPV prevalence, and these outcomes are influenced by infection persistence, disease progression, screening practices, and diagnostic processes in addition to transmission.^57^ As longer-term post-vaccination evidence accumulates in low- and middle-income countries, future work could expand our model by incorporating data from those settings to evaluate how indirect protection varies across different population settings.^64,65^ In addition, while most existing studies focus on the indirect benefits of female immunisation, evidence regarding the health impact of HPV vaccination on males has expanded rapidly in recent years.^66^ Future research should examine indirect effects among boys and men and assess how these effects differ between female-only and gender-neutral vaccination programmes.

In conclusion, our analysis of real-world post-vaccination data suggests that population-level indirect protection against HPV16/18 develops gradually following vaccine introduction and varies across age and vaccination-eligibility groups. These findings provide explicit characterisation of indirect protection as HPV vaccination programmes mature and can complement transmission-dynamic models used to evaluate the population-level effects of HPV vaccination.

## Contributors

Conceptualisation: RW and MJ. Data collection, analysis, and original manuscript drafting: RW. Supervision: MJ. All authors contributed to the results interpretation, critical review, and manuscript revision, and have approved the final version. All authors had full access to the data.

## Data sharing statement

All analysis code is available at https://github.com/RZJosieW/HPV-vaccine--Indirect-effect.

## Data Availability

All analysis code and data are available at https://github.com/RZJosieW/HPV-vaccine--Indirect-effect.

https://github.com/RZJosieW/HPV-vaccine--Indirect-effect

## Acknowledgements

We thank Kiesha Prem for helpful comments on the study design. This study received no specific funding.

