## Appendix 1 for "Population-level indirect effects of HPV vaccination in high-income settings: a Bayesian modelling study"

　　5.1 Model specification ......................................................................... 10–12
　　5.2 Model diagnostics ............................................................................ 13–14
　　5.3 Posterior distributions and parameter estimates .............................. 15–18

**6. Sensitivity analyses ............................................................................... 19**
　　6.1 Model results excluding the US and UK ............................................ 19–20
　　6.2 Model results for the US and UK only ............................................... 21–22

**7. Study-level random-effect estimates ................................................... 23**

**8. HPV vaccination programme schedules ........................................ 24–33**

**References ........................................................................................... 34–35**

1. **Rapid systematic review**
   1.1 **Method**

To extend the latest evidence from population-level studies of HPV vaccination, we conducted a rapid systematic review by following the search and reporting procedures outlined in the Preferred Reporting Items for Systematic Reviews and Meta-Analyses (PRISMA) guidelines (Supplementary Figure 1). We searched published literature through PubMed between October 11, 2018, and July 20, 2025. We used the pre-defined search strategy from two previous systematic reviews to identify relevant literature.^1,2^

We modified the inclusion criteria based on the previous studies to better fit with the objectives of our study.^1,2^ After removing duplicate articles, we screened and identified studies conducted in high-income countries, comparing the frequency (prevalence or incidence) of genital HPV infections between pre-vaccination and post-vaccination periods in the general population and using the same population sources and recruitment methods before and after vaccination.

We excluded any study if: 1) the study was a review or modelling study, 2) the reported prevalence or incidence of genital HPV infection was only in males, 3) HPV vaccination was administered as part of a randomised trial, or 4) data for the pre-vaccination period were not available.

During this rapid systematic review, we did not critically assess the quality of the identified studies.

RW independently conducted the literature search and study screening.

1.2 **Results**

We identified a total of 2,411 articles using the search term. After removing two duplicate articles, 2,409 articles remained for title and abstract screening. Based on the study inclusion and exclusion criteria, 2,336 non-eligible studies were excluded.

We then downloaded the PDF versions of the remaining 73 articles from the publishers’ websites for full-text assessment. Of these, 61 articles were excluded after full-text review. In total, 12 eligible studies were included.

**Identification of studies via databases and registers**

Electronic Database searches:

PubMed

(n=2411)

**Identification**

Duplicate records removed	(n=2)

Exclude (n=2336)

Mathematical modelling (n=35)

Review papers (n=197)

No descriptions of population-level effects of human papillomavirus vaccination (n=1913)

Non-high-income countries (n=191)

Title and abstracts screened

(n= 2409)

**Screening**

Exclude (n=61)

Duplicate (n=2)

Study on men only (n=4)

Endpoint does not include HPV infection (n=46)

Compared unvaccinated and vaccinated women only (n=9)

Full-text articles assessed for eligibility

(n=73)

Studies included in this analysis

(n=12)

**Included**

**Supplementary Figure 1. PRISMA flow diagram.**

2. **Data preparation**
   2.1 **Conversion of prevalence ratio to age-specific prevalence in** **Purriños-Hermida et al.**

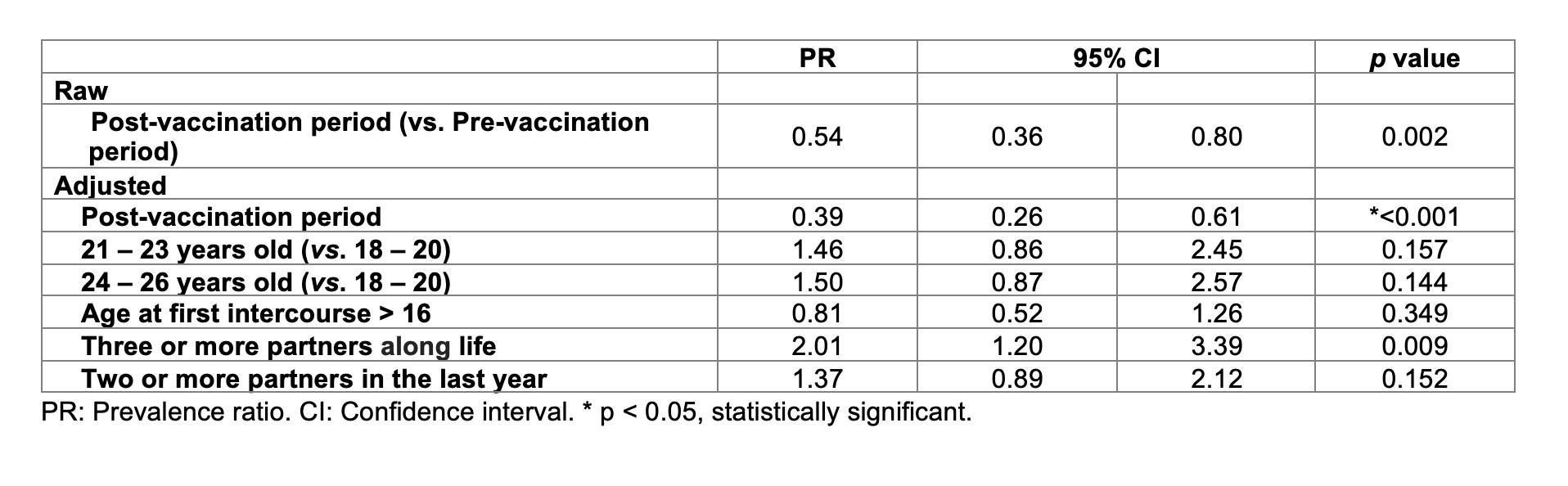

**Supplementary Figure 2. Result table for the Purriños-Hermida et al.**

In Purriños-Hermida et al.^3^, the authors report only the overall prevalence ratio (PR) for HR-HPV 16/18 and its 95% confidence interval (CI) in vaccinated and unvaccinated women in the post-vaccination period versus women in the pre-vaccination period (Supporting information S4 Table of Purriños-Hermida et al.^3^). To estimate the age-specific prevalence in both the post- and pre-vaccination periods, we combined the overall prevalence (5·2%), the adjusted age-specific PRs from supporting information S4 Table, and the distribution of participants across age groups from Table 2 of Purriños-Hermida et al.^3^.

### Age-specific prevalence in the post-vaccination period

From Table 2, the number of individuals in the post-vaccination period is:

Age 18–20: 238, Age 21–23: 315, Age 24–26: 192

As the total number of participants in the post-vaccination period is 745, the sample weights are:

$w_{1}=\frac{238}{745}\approx0\text{·}319,w_{2}=\frac{315}{745}\approx0\text{·}423,w_{3}=\frac{192}{745}\approx0\text{·}258$

Let the prevalence for the 18–20 age group be p. Using the adjusted PRs:

$\mathrm{Prevalence}_{21-23}=1\text{·}46\cdot p$

$\mathrm{Prevalence}_{24-26}=1\text{·}50\cdot p$

The total prevalence is the weighted average:

$\text{Total Post-vaccination Prevalence}=p\cdot\left( w_{1}+1\text{·}46\cdot w_{2}+1\text{·}50\cdot w_{3} \right)$

$5\text{·}2\%=p\cdot\left( 0\text{·}319+1\text{·}46\cdot0\text{·}423+1\text{·}50\cdot0\text{·}258 \right)$

$=p\cdot\left( 0\text{·}319+0\text{·}617+0\text{·}387 \right)=p\cdot1\text{·}323$

$\Rightarrow p=\frac{5\text{·}2\%}{1\text{·}323}\approx3\text{·}93\%$

The estimated age-specific post-vaccination prevalences are:

$\mathrm{Prevalence}_{\text{18–20}}=\text{3}\text{·}\text{93\%}$

$\mathrm{Prevalence}_{\text{21–23}}=\text{1.46 · 3.93\% ≈ 5}\text{·}\text{74\%}$

$\mathrm{Prevalence}_{\text{24–26}}=\text{1.50 · 3.93\% ≈ 5}\text{·}\text{90\%}$

### Age-specific prevalence in the pre-vaccination period

From Table 2, the number of individuals in the pre-vaccination period is:

Age 18–20: 188, Age 21–23: 161, Age 24–26: 174

As the total number of participants in the pre-vaccination period is 523, the sample weights are:

$w_{1}=\frac{188}{523}\approx0\text{·}360,w_{2}=\frac{161}{523}\approx0\text{·}308,w_{3}=\frac{174}{523}\approx0\text{·}333$

Let the prevalence for the 18–20 age group in the pre-vaccination period be q. With a similar approach:

$\text{Total Pre-vaccination Prevalence}=q\cdot\left( w_{1}+1\text{·}46\cdot w_{2}+1\text{·}50\cdot w_{3} \right)$

$9\text{·}8\%=q\cdot\left( 0\text{·}360+1\text{·}46\cdot0\text{·}308+1\text{·}50\cdot0\text{·}333 \right)$

$\Rightarrow q=\frac{9\cdot8\%}{1\cdot309}\approx\text{7}\text{·}\text{49\%}$

The estimated age-specific pre-vaccination prevalences are:

$\mathrm{Prevalence}_{\text{18–20}}=\text{7}\text{·}\text{49\%}$

$\mathrm{Prevalence}_{\text{21–23}}\approx\text{10}\text{·}\text{94\%}$

$\mathrm{Prevalence}_{\text{24–26}}\approx\text{11}\text{·}\text{24\%}$

2.2 **Interpolation of age-specific HPV prevalence**

Many studies provide HPV prevalence in large age ranges. To estimate age-specific prevalence for single years of age, we applied exponentially damped models to data in the age ranges provided. These are a flexible family of models that capture the shape of the age distribution of HPV prevalence in many high-income countries. This is similar to the approach used in a previous HPV model:^4^

$f\left( x \right)=B+\left[ A\left( x-x_{0} \right)^{n}-B \right]\times e^{-C\left( x-x_{0} \right)},n\in\left\{ 1.0,1.1,\ldots,3.0 \right\}$	(1)

Here, x denotes age, f(x) is the estimated HPV prevalence at age x, and x₀ is the reference age used to centre the curve. The parameters A, B, and C determine the shape of the fitted prevalence curve, while n controls the polynomial component of the age-specific relationship.

For age group i, let xᵢ denote the midpoint of the reported age interval, Nᵢ the corresponding sample size, and yᵢ the number of individuals with HPV16/18 infection. We assumed that the number of infected individuals followed a binomial distribution. For each candidate value of n, the parameters A, B, and C were estimated by maximising the binomial likelihood, equivalently by minimising the negative binomial log-likelihood:

$\overset{^}{A},\overset{^}{B},\overset{^}{C}=\underset{A,B,C}{\text{argmin}}\left\{ -\sum_{i=1}^{K} \left[ y_{i}\log f\left( x_{i} \right)+\left( N_{i}-y_{i} \right)\log\left( 1-f\left( x_{i} \right) \right) \right] \right\}$	(2)

The Broyden–Fletcher–Goldfarb–Shanno (BFGS) algorithm was applied using the R function *optim* to obtain maximum likelihood estimates of the parameter values.^5^ The model was fitted for values of n ranging from 1·0 to 3·0 in increments of 0·1. For each value of n, the parameters A, B, and C were estimated separately. The value of n with the lowest optimized negative log-likelihood under the binomial model was used for interpolation. Markov Chain Monte Carlo (MCMC) methods were used to estimate the credible intervals for the predicted prevalence, using the parameter estimates obtained by BFGS optimization as the reference values for setting priors for A, B, and C.

2.3 **Reconstruction of age-specific vaccine coverage**

### (1) Vaccine coverage is reported in the article

If the article reported vaccine coverage in the post-vaccination period, we use the values provided directly.

### (2) Vaccine coverage is not reported in the article

If the article did not report vaccine coverage in the post-vaccination period, we use data from government reports or websites in the same country to calculate vaccine coverage by age and year.

Let A = [aij] be a matrix of vaccine coverage by age and calendar year, where the row index i corresponds to age and the column index j corresponds to calendar year. For individuals aged a in year t, their birth year is b = t − a. We define the vaccine coverage for cohort b as vt−a. Then each matrix entry is equal to:

$a_{ij}=v_{t-a}$

As the national immunisation programme in most countries is only eligible for specific cohorts at a given time, all entries along the diagonal where j − i is constant (i.e., the same birth cohort) are assigned the same value (Supplementary Table 1):

$a_{ij}=a_{i+1,j+1}=a_{i+2,j+2}=\cdots=v_{t-a}$

| **Age \ Year** | **2012** | **2013** | **2014** | **2015** | **2016** | **2017** | **2018** |
| --- | --- | --- | --- | --- | --- | --- | --- |
| **13** | v₁₉₉₉ | v₂₀₀₀ | v₂₀₀₁ | v₂₀₀₂ | v₂₀₀₃ | v₂₀₀₄ | v₂₀₀₅ |
| **14** | v₁₉₉₈ | v₁₉₉₉ | v₂₀₀₀ | v₂₀₀₁ | v₂₀₀₂ | v₂₀₀₃ | v₂₀₀₄ |
| **15** | v₁₉₉₇ | v₁₉₉₈ | v₁₉₉₉ | v₂₀₀₀ | v₂₀₀₁ | v₂₀₀₂ | v₂₀₀₃ |
| **16** | v₁₉₉₆ | v₁₉₉₇ | v₁₉₉₈ | v₁₉₉₉ | v₂₀₀₀ | v₂₀₀₁ | v₂₀₀₂ |

**Supplementary Table 1. Reconstruction of HPV vaccine coverage by age, calendar year, and birth cohort.**

3. **Propagation of uncertainty**

Given the varying availability of data across studies, we consider two scenarios to propagate uncertainty in the estimation of indirect effects. As only a few studies report confidence intervals for vaccine coverage, we treat all vaccine coverage values as fixed data points in both scenarios. Both scenarios use a Monte Carlo approach. We assumed VEITT = 1 (100%) in both scenarios.

### **(**1) Scenario 1

If a study provides HPV prevalence across multiple age groups for both pre- and post-vaccination periods, the uncertainty in the indirect effect was propagated through posterior sampling.

The estimated pre- and post-vaccination age-specific prevalence and their corresponding credible intervals were obtained through interpolation using the method described in Section 2. For each age a and corresponding vaccine coverage v(a), we drew samples from the posterior distributions of the pre- and post-vaccination age-specific prevalence estimates. For each posterior draw s, we then computed the total, direct, and indirect effects as:

$\mathrm{VE}_{\text{mITT}}^{\left( s \right)}\left( a \right)=\mathrm{VE}_{\text{ITT}}\left[ 1-p_{\text{post}}^{\left( s \right)}\left( a \right) \right]$	(3)

$\mathrm{Total}^{\left( s \right)}\left( a \right)=p_{\text{pre}}^{\left( s \right)}\left( a \right)-p_{\text{post}}^{\left( s \right)}\left( a \right)$	(4)

$\mathrm{Direct}^{\left( s \right)}\left( a \right)=p_{\text{pre}}^{\left( s \right)}\left( a \right)v\left( a \right)\mathrm{VE}_{\text{mITT}}^{\left( s \right)}\left( a \right)$	(5)

$\mathrm{Indirect}^{\left( s \right)}\left( a \right)=\mathrm{Total}^{\left( s \right)}\left( a \right)-\mathrm{Direct}^{\left( s \right)}\left( a \right)$	(6)

$\mathrm{Indirect}^{\left( s \right)}\left( a \right)=p_{\text{pre}}^{\left( s \right)}\left( a \right)-p_{\text{post}}^{\left( s \right)}\left( a \right)-p_{\text{pre}}^{\left( s \right)}\left( a \right)v\left( a \right)\mathrm{VE}_{\text{ITT}}\left[ 1-p_{\text{post}}^{\left( s \right)}\left( a \right) \right]$

Here, $p_{\text{pre}}^{\left( s \right)}\left( a \right)$ and $p_{\text{post}}^{\left( s \right)}\left( a \right)$ represent the posterior-predicted prevalence in the pre- and post-vaccination periods. Using 4,000 posterior draws, we calculated the posterior mean, standard deviation, and 95% credible interval (CrI) of the estimated indirect effect for each age.

Hence this scenario captures uncertainty in the underlying prevalence curve, conditional on the observed data (including sampling uncertainty through the binomial likelihood) and the interpolating model used.

### (2) Scenario 2

For studies that provided only point estimates of HPV prevalence for the pre- and/or post-vaccination periods, we constructed distributions for the corresponding prevalence estimates and propagated this uncertainty using Monte Carlo sampling.

For each age a, we assume both estimated pre- and post-vaccination prevalence follow the normal distribution:

$p_{\text{pre}}^{\left( s \right)}\left( a \right)\sim N\left( \overset{^}{p_{\text{pre}}}\left( a \right),{\mathrm{SE}_{\text{pre}}}^{2}\left( a \right) \right),p_{\text{post}}^{\left( s \right)}\left( a \right)\sim N\left( \overset{^}{p_{\text{post}}}\left( a \right),{\mathrm{SE}_{\text{post}}}^{2}\left( a \right) \right)$

where SE(p̂) was estimated using	$\mathrm{SE}\left( \overset{^}{p} \right)=\sqrt{\frac{\overset{^}{p}\left( 1-\overset{^}{p} \right)}{n}}$

where n is the sample size and p̂ is the HPV prevalence.

Given vaccine coverage v(a), for each Monte Carlo draw s, we computed:

$\mathrm{Total}^{\left( s \right)}\left( a \right)=p_{\text{pre}}^{\left( s \right)}\left( a \right)-p_{\text{post}}^{\left( s \right)}\left( a \right)$

$\mathrm{Direct}^{\left( s \right)}\left( a \right)=p_{\text{pre}}^{\left( s \right)}\left( a \right)v\left( a \right)\mathrm{VE}_{\text{ITT}}\left[ 1-p_{\text{post}}^{\left( s \right)}\left( a \right) \right]$

$\mathrm{Indirect}^{\left( s \right)}\left( a \right)=\mathrm{Total}^{\left( s \right)}\left( a \right)-\mathrm{Direct}^{\left( s \right)}\left( a \right)$

Using 4,000 Monte Carlo draws, we calculated the mean, standard deviation, and 95% uncertainty interval of the estimated indirect effect for each age.

Hence this scenario captures approximate binomial sampling uncertainty in directly observed prevalence estimate

4. **Descriptive summary of data extracted from the review**

The distributions of the main variables included in the analysis are presented in Supplementary Figure 3. Estimated indirect effects were predominantly positive, while vaccine coverage varied widely across observations. Age at effect assessment was concentrated among adolescents and young adults. The dataset included observations from both early and later post-vaccination periods, extending to 12·5 years after vaccine introduction.
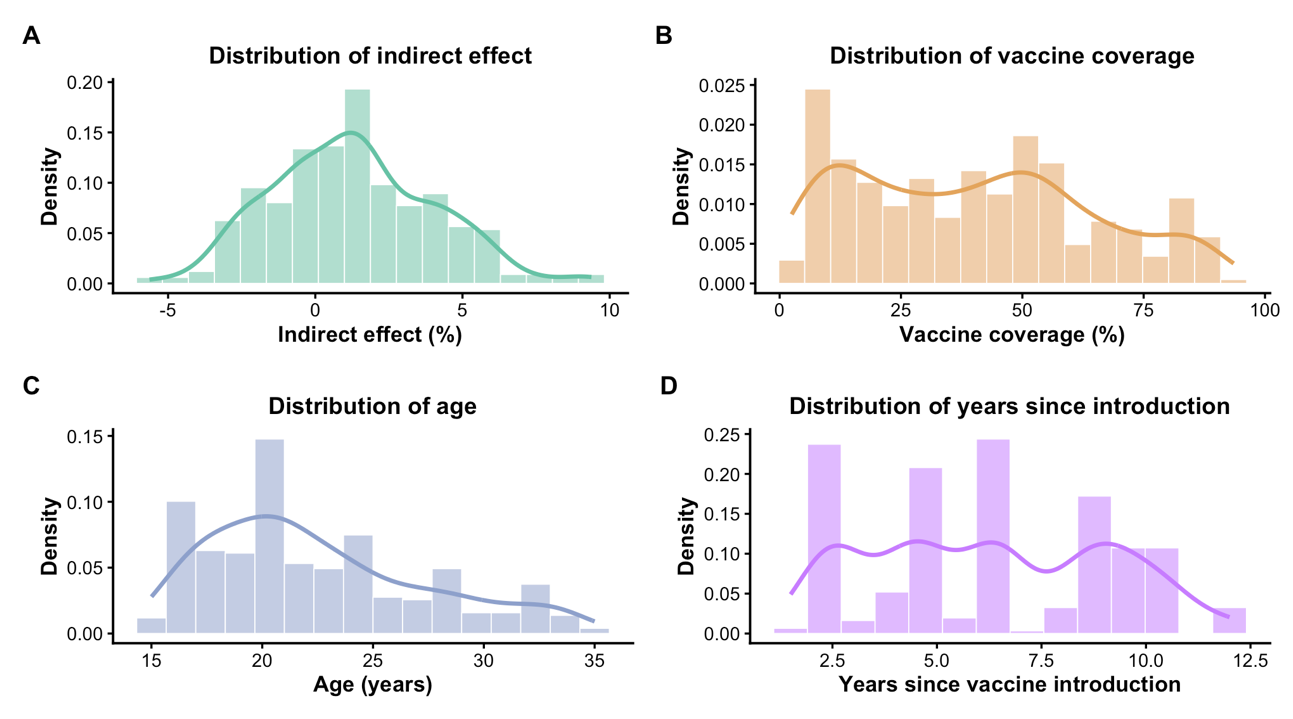
**Supplementary Figure 3. Distribution of key variables.** Histograms and kernel density curves showing the distributions of (A) indirect effect (%), (B) vaccine coverage (%), (C) age at effect assessment, and (D) years since vaccine introduction.

Supplementary Figure 4 showed the distribution of age across the routine, catch-up, and non-targeted groups. The routine group consisted mainly of younger individuals (15–24 years), with most observations clustered around late adolescence (15-18 years). The catch-up group showed a broader age distribution, ranging from approximately 15 to 34 years. In contrast, the non-targeted group included older individuals than the routine and catch-up groups, with ages ranging from 18 to 54 years.

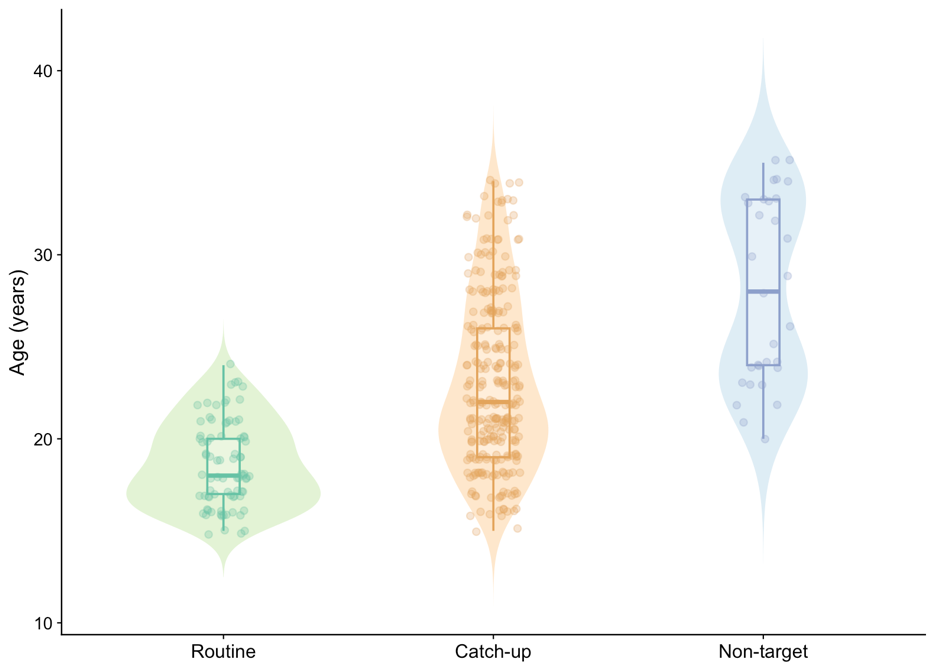

**Supplementary Figure 4. Distribution of age at effect assessment across vaccination-eligibility groups.**

As shown in Supplementary Figure 5, the catch-up group comprised the largest percentage of observations included in the analysis (70·6%), followed by the routine group (21·3%) and the non-targeted group (8·1%).
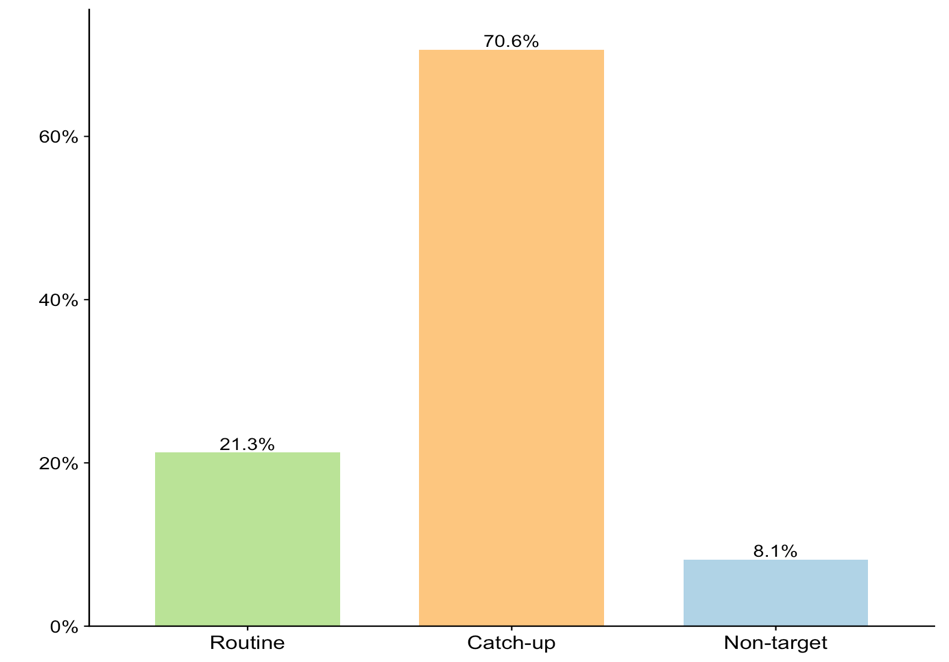

**Supplementary Figure 5. Distribution of observations by vaccination-eligibility group.**

5. **Bayesian hierarchical model**

We used a Bayesian hierarchical model as the primary analysis to characterise the associations between the estimated indirect effect and vaccine coverage, time since vaccine introduction, and age at effect assessment. We modelled the estimated indirect effect as the outcome using a Student’s t likelihood. The standard error of each indirect effect estimate was also included in the model to account for its uncertainty. Vaccination-eligibility group (routine, catch-up, or non-targeted) was included as a categorical fixed effect. Non-linear associations with vaccine coverage, time since vaccine introduction, and age were modelled using smooth functions. For each predictor, we included both an overall smooth and vaccination-eligibility-group-specific smooths, allowing the shape of each association to differ across the three groups. Random intercepts were included for each country-by-vaccination-eligibility group and for each study to account for heterogeneity across countries, vaccination groups, and studies.

### **5.1 Model details**

Let Yc,j,z,i denote the estimated indirect effect for observation i, from country c, vaccination-eligibility group j, and study z. We assumed a Student’s t likelihood:

$Y_{c,j,z,i}\sim t_{\nu}\left( \mu_{c,j,z,i},\sqrt{{\mathrm{SE}_{i}}^{2}+\sigma^{2}} \right)$

where SEi is the standard error associated with the estimated indirect effect and σ represents additional residual variation.

The expected indirect effect was modelled as:

$\begin{aligned} \mu_{c,j,z,i}=\alpha+\beta_{1}I\left( j=\text{catchup} \right)+\beta_{2}I\left( j=\text{nontarget} \right) \\ +f_{1}\left( \mathrm{Coverage}_{i} \right)+f_{2}\left( \mathrm{Time}_{i} \right)+f_{3}\left( \mathrm{Age}_{i} \right) \\ +f_{1,j}\left( \mathrm{Coverage}_{i} \right)+f_{2,j}\left( \mathrm{Time}_{i} \right)+f_{3,j}\left( \mathrm{Age}_{i} \right) \\ +u_{c,j}+v_{z}. \end{aligned}$

The routine-vaccination group was used as the reference category for the parametric vaccination-eligibility-group effect. Separate group-specific smooth terms were estimated for all three vaccination-eligibility groups. The functions f1(·), f2(·), and f3(·) represent the overall non-linear associations with vaccine coverage, time since vaccine introduction, and age at effect assessment, respectively. The functions f1,j(·), f2,j(·), and f3,j(·) represent additional vaccination-eligibility-group-specific smooth terms.

Thin-plate regression splines were used for all smooth functions. The basis dimensions for the overall smooths were k = 8 for vaccine coverage and k = 6 for both time since vaccine introduction and age at effect assessment. For the vaccination-eligibility-group-specific smooths, the basis dimensions were k = 6 for vaccine coverage and k = 4 for both time since vaccine introduction and age at effect assessment.

The random intercept uc,j represents variation between country-by-vaccination-eligibility-group combinations, allowing partial pooling across observations within the hierarchical model. The study-level random intercept vz accounts for residual heterogeneity between studies after accounting for the modelled predictors and country-by-vaccination-eligibility effects. Definitions of the variables and parameters used in the model are provided in Supplementary Table 2.

Weakly informative prior distributions were used for the model parameters. The intercept and fixed-effect coefficients were assigned Student-t priors with 3 degrees of freedom, location 0, and scale 5. The degrees-of-freedom parameter of the Student’s t likelihood was assigned a Gamma (2, 0·1) prior. Standard deviations of the smooth terms were assigned Exponential (0·5) priors, and standard deviations of the country-by-vaccination-eligibility-group and study-level random effects were assigned Exponential (1·5) priors. The complete prior assumption is summarised in Supplementary Table 3.

| **Variable** | **Description** |
| --- | --- |
| $Y_{c,j,z,i}$ | Estimated indirect effect for observation i in country c, group j, and study z |
| $\mu_{c,j,z,i}$ | Expected indirect effect for the corresponding observation |
| ${SE}_{i}$ | Standard error of the estimated indirect effect for observation i |
| $i$ | Observation index, i = 1, …, N |
| $c$ | Country index, c = 1, …, C |
| $j$ | Group index: 1 = routine; 2 = catch-up; 3 = non-targeted |
| $z$ | Study index, z = 1, …, Z |
| $h$ | Predictor index: 1 = coverage; 2 = time since introduction; 3 = age |
| $k$ | Index of a reported spline coefficient within a smooth term |
| ${Coverage}_{i}$ | Vaccine coverage for observation i |
| ${Time}_{i}$ | Time since vaccine introduction for observation i |
| ${Age}_{i}$ | Age at effect assessment for observation i |
| $\alpha$ | Intercept under the fitted brms parameterisation |
| $\beta_{2}$ | Catch-up versus routine parametric group coefficient |
| $\beta_{3}$ | Non-targeted versus routine parametric group coefficient |
| $f_{h}$ | Overall smooth function for predictor h |
| $f_{h,j}$ | Additional group-specific smooth function for predictor h in group j |
| $\theta_{h,k}$ | kth reported spline coefficient associated with overall smooth fₕ |
| $\theta_{h,j,k}$ | kth reported spline coefficient associated with group-specific smooth fₕ,ⱼ |
| $u_{c,j}$ | Country-by-group random intercept |
| $v_{z}$ | Study random intercept |
| $\sigma_{f,h}$ | SD parameter of the penalised component of overall smooth fₕ |
| $\sigma_{f,h,j}$ | SD parameter of the penalised component of group-specific smooth fₕ,ⱼ |
| $\sigma_{u}$ | SD of country-by-group random intercepts |
| $\sigma_{v}$ | SD of study random intercepts |
| $\sigma$ | Additional residual scale parameter of the Student’s t likelihood |
| $\nu$ | Degrees of freedom of the Student’s t likelihood |

**Supplementary Table 2. Definitions of variables and parameters in the Bayesian hierarchical model.**

| **Parameter** | **Prior** |
| --- | --- |
| $\alpha$ | Student-t(3, 0, 5) |
| $\beta_{2}$ | Student-t(3, 0, 5) |
| $\beta_{3}$ | Student-t(3, 0, 5) |
| $\nu$ | Gamma(2, 0·1) |
| $\sigma_{f,h}$ | Exponential(0·5) |
| $\sigma_{f,h,j}$ | Exponential(0·5) |
| $\sigma_{u}$ | Exponential(1·5) |
| $\sigma_{v}$ | Exponential(1·5) |

**Supplementary Table 3. Prior distributions specified for the Bayesian hierarchical model.**

**5.2 Model diagnostics**

We used multiple diagnostic approaches (trace plots, posterior predictive checks, R-hat statistics, and effective sample size diagnostics) to assess model convergence and stability of the model outputs.

The R-hat values for all parameters ranged from 0·9998 to 1·002. Based on 12,000 post-warmup draws, the effective sample size ratios ranged from 0·10 to 0·90 (median = 0·57). The trace plots in Supplementary Figure 6 showed good chain mixing and model convergence. The posterior predictive distributions overlapped with the observed distribution in Supplementary Figure 7, indicating that the model can capture the distributional characteristics of the observed data.

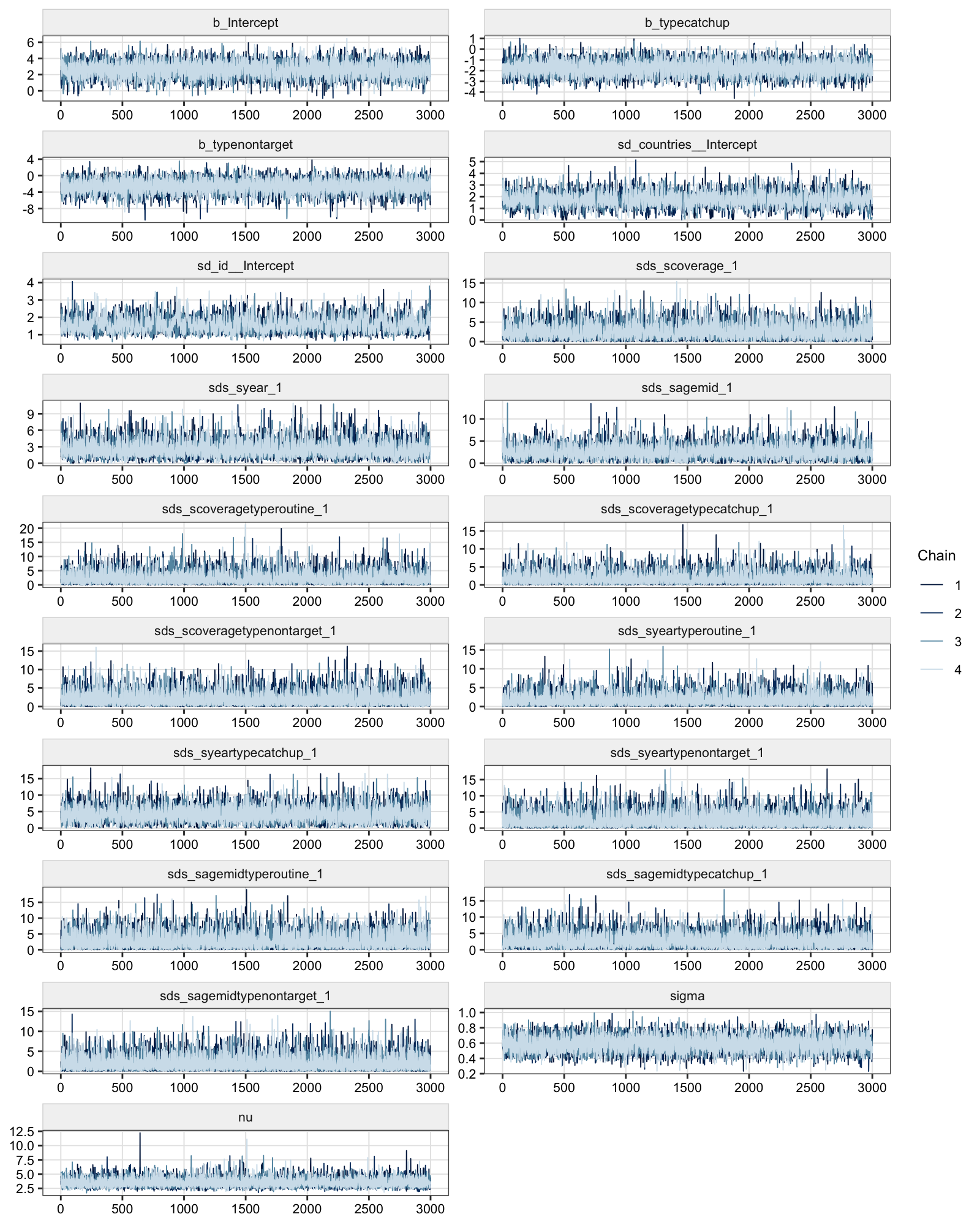

**Supplementary Figure 6. MCMC trace plots.** The plots showed the iteration trajectories of the four independent Markov chains (represented by different shades of blue) for the main model parameters.

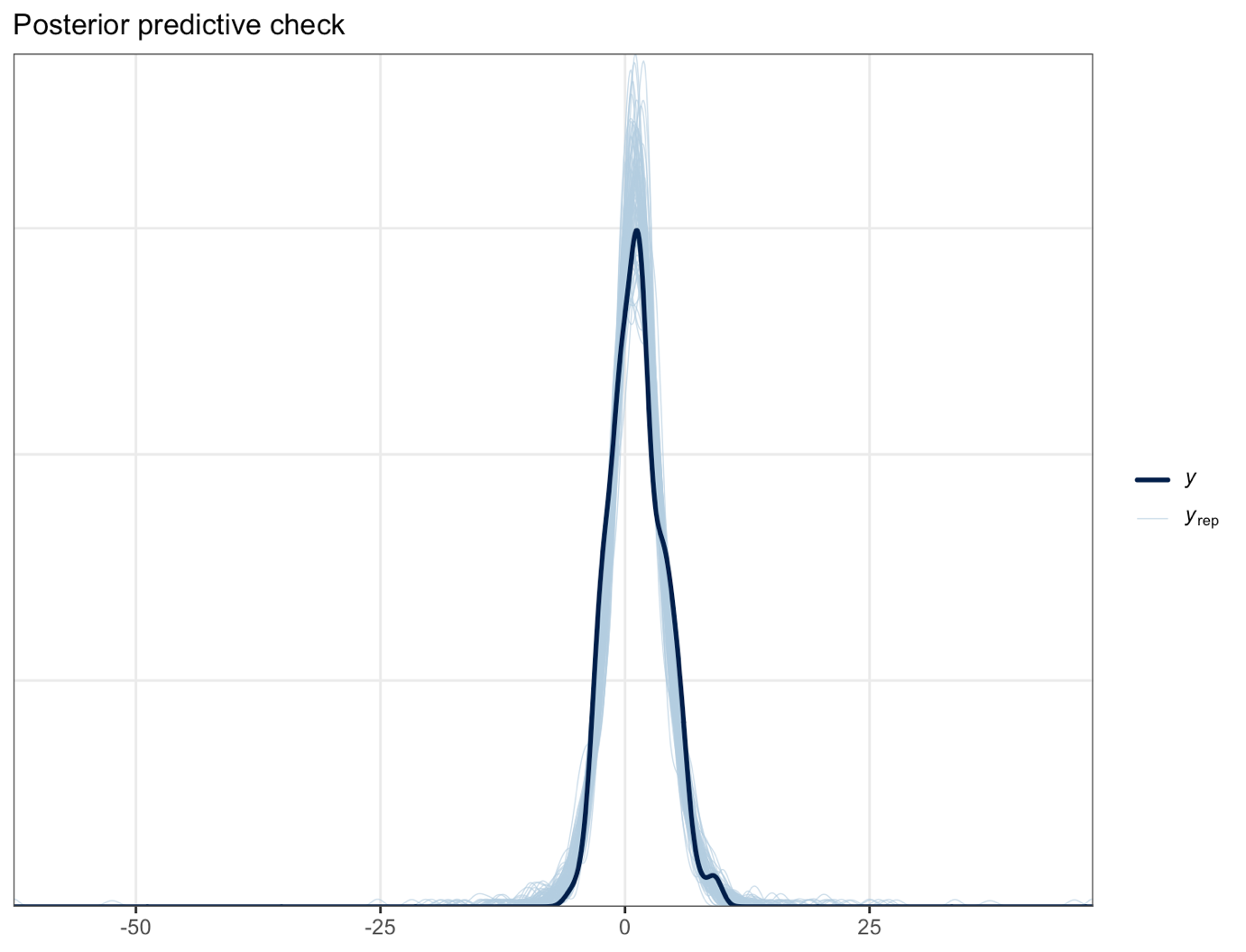

**Supplementary Figure 7. Posterior predictive check.** Posterior predictive check comparing the observed distribution (dark blue curve) with replicated posterior predictive distributions (light blue curve) generated from the model.

**5.3 Posterior distributions and parameter summaries**

Posterior distributions are shown for the parametric coefficients, spline coefficients, and hyperparameters, with corresponding posterior means, standard deviations, and 95% credible intervals reported in the tables. In each plot, the vertical red line indicates the posterior mean, and the horizontal red line indicates the 95% credible interval.

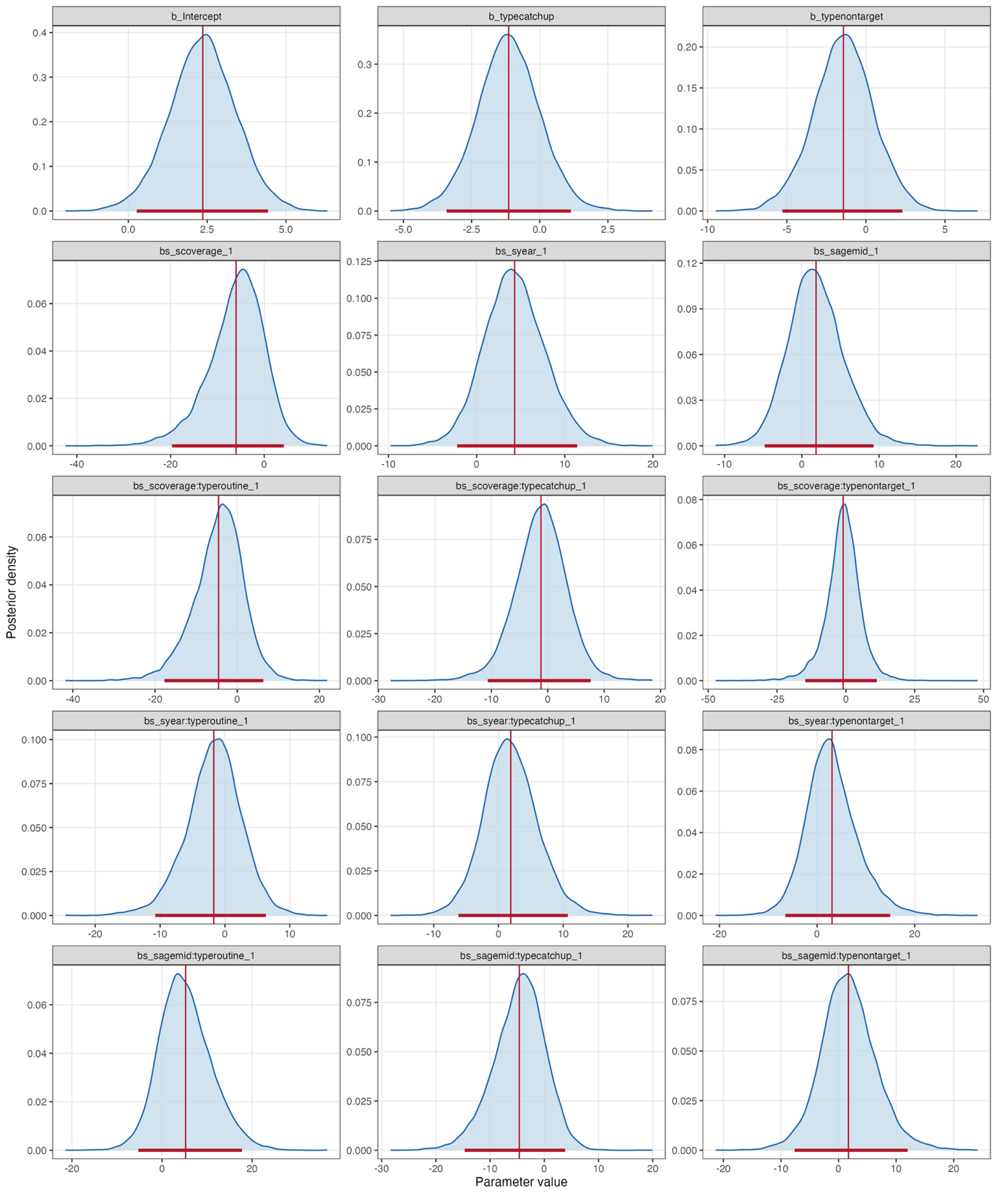

**Supplementary Figure 8. Posterior distributions of the model coefficients, including spline coefficients.**

| **Model component** | **Parameter** | **Posterior mean** | **Posterior SD** | **95% CrI** |
| --- | --- | --- | --- | --- |
| Intercept | *α* | 2·362 | 1·045 | [0·267, 4·430] |
| Catch-up vs routine: parametric group term | *β*_2_ | -1·140 | 1·150 | [-3·418, 1·145] |
| Non-targeted vs routine: parametric group term | *β*_3_ | -1·420 | 1·908 | [-5·279, 2·309] |
| Spline coefficient: coverage smooth (overall) | *θ*_1,1_ | -6·007 | 6·001 | [-19·695, 4·194] |
| Spline coefficient: time since introduction smooth (overall) | *θ*_2,1_ | 4·315 | 3·449 | [-2·207, 11·409] |
| Spline coefficient: age smooth (overall) | *θ*_3,1_ | 1·851 | 3·607 | [-4·788, 9·320] |
| Spline coefficient: coverage smooth (routine) | *θ*_1,1,1_ | -4·545 | 6·065 | [-17·709, 6·351] |
| Spline coefficient: coverage smooth (catch-up) | *θ*_1,2,1_ | -1·201 | 4·630 | [-10·629, 7·629] |
| Spline coefficient: coverage smooth (non-targeted) | *θ*_1,3,1_ | -1·002 | 6·401 | [-14·761, 11·211] |
| Spline coefficient: time since introduction smooth (routine) | *θ*_2,1,1_ | -1·718 | 4·295 | [-10·743, 6·308] |
| Spline coefficient: time since introduction smooth (catch-up) | *θ*_2,2,1_ | 1·899 | 4·245 | [-6·184, 10·724] |
| Spline coefficient: time since introduction smooth (non-targeted) | *θ*_2,3,1_ | 3·067 | 5·369 | [-6·493, 14·976] |
| Spline coefficient: age smooth (routine) | *θ*_3,1,1_ | 5·235 | 5·839 | [-5·270, 17·756] |
| Spline coefficient: age smooth (catch-up) | *θ*_3,2,1_ | -4·620 | 4·734 | [-14·701, 3·842] |
| Spline coefficient: age smooth (non-targeted) | *θ*_3,3,1_ | 1·721 | 4·869 | [-7·637, 12·002] |

**Supplementary Table 4. Posterior summaries of population-level and spline coefficients**

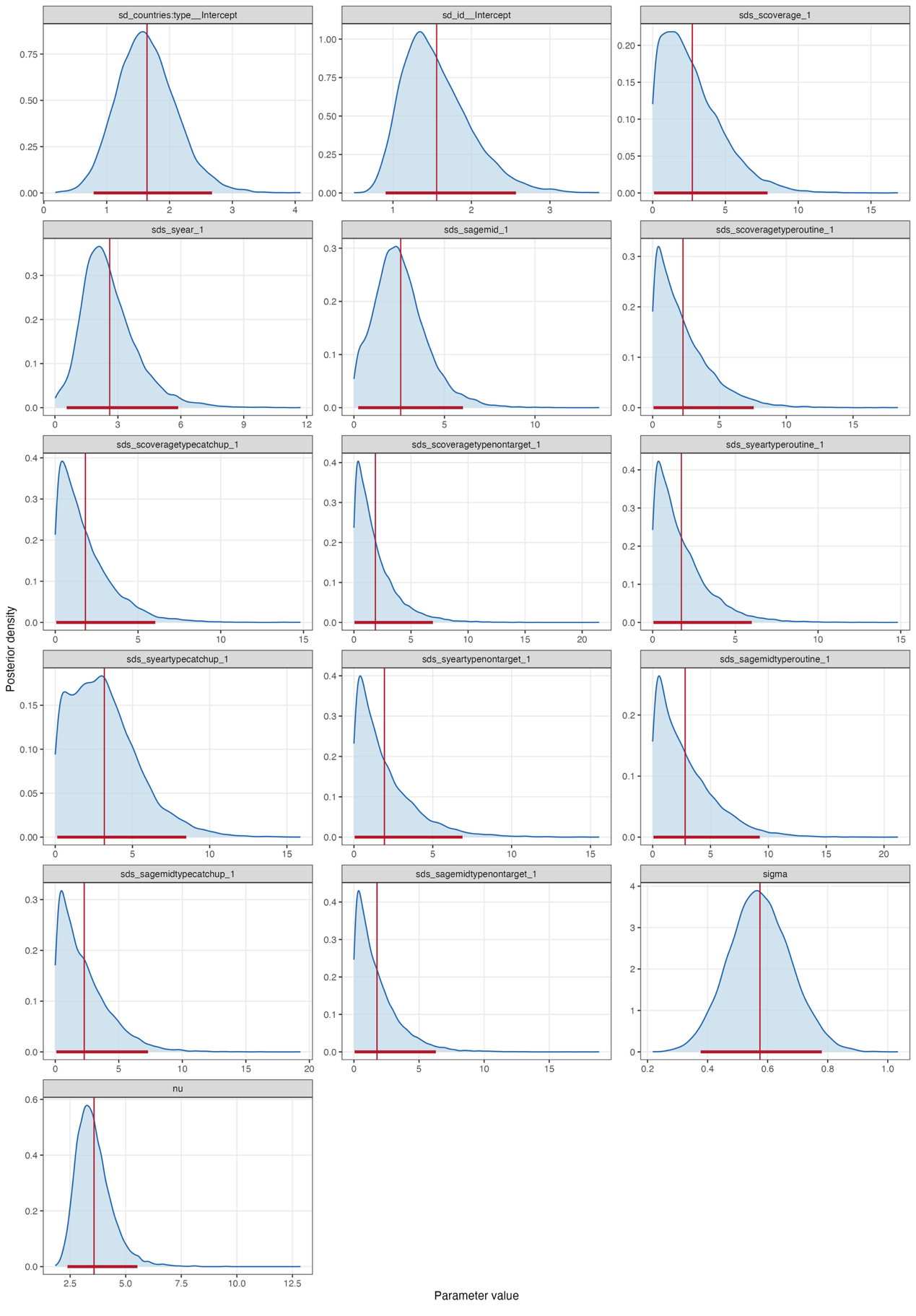

**Supplementary Figure 9. Posterior distributions of the model hyperparameters.**

| **Model component** | **Parameter** | **Posterior mean** | **Posterior SD** | **95% CrI** |
| --- | --- | --- | --- | --- |
| SD of country-by-group random intercepts | *σ*_u_ | 1·642 | 0·477 | [0·793, 2·676] |
| SD of study random intercepts | *σ*_v_ | 1·557 | 0·431 | [0·906, 2·568] |
| Spline SD: coverage smooth (overall) | *σ*_f,1_ | 2·725 | 2·092 | [0·093, 7·893] |
| Spline SD: time since introduction smooth (overall) | *σ*_f,2_ | 2·608 | 1·341 | [0·558, 5·876] |
| Spline SD: age smooth (overall) | *σ*_f,3_ | 2·582 | 1·467 | [0·236, 6·025] |
| Spline SD: coverage smooth (routine) | *σ*_f,1,1_ | 2·271 | 2·083 | [0·064, 7·569] |
| Spline SD: coverage smooth (catch-up) | *σ*_f,1,2_ | 1·821 | 1·656 | [0·061, 6·045] |
| Spline SD: coverage smooth (non-targeted) | *σ*_f,1,3_ | 1·877 | 1·852 | [0·048, 6·920] |
| Spline SD: time since introduction smooth (routine) | *σ*_f,2,1_ | 1·737 | 1·622 | [0·048, 5·993] |
| Spline SD: time since introduction smooth (catch-up) | *σ*_f,2,2_ | 3·178 | 2·222 | [0·129, 8·484] |
| Spline SD: time since introduction smooth (non-targeted) | *σ*_f,2,3_ | 1·933 | 1·877 | [0·048, 6·884] |
| Spline SD: age smooth (routine) | *σ*_f,3,1_ | 2·807 | 2·528 | [0·069, 9·260] |
| Spline SD: age smooth (catch-up) | *σ*_f,3,2_ | 2·281 | 1·995 | [0·083, 7·301] |
| Spline SD: age smooth (non-targeted) | *σ*_f,3,3_ | 1·771 | 1·714 | [0·052, 6·289] |
| Additional residual scale | *σ* | 0·574 | 0·103 | [0·377, 0·780] |
| Student-t degrees of freedom | *ν* | 3·575 | 0·809 | [2·382, 5·528] |

**Supplementary Table 5. Posterior summaries of model hyperparameters**

6. **Sensitivity analyses**

The United States (US) and the United Kingdom (UK) accounted for 67·7% of the observations in the overall dataset. We therefore conducted sensitivity analyses to assess whether the primary results were influenced by the large contribution of these two countries. Two scenarios were considered: (1) excluding all observations from the US and UK, and (2) restricting the analysis to observations from the US and UK.

**6.1 Model Results Without US and UK**

After excluding observations from the US and UK, the overall associations remained broadly similar to those observed in the primary analysis, although some differences were observed in the shapes of the fitted relationships (Supplementary Figure 10). The association with vaccine coverage showed an initial increase followed by a gradual decline at higher coverage levels. The association with time since vaccine introduction remained similar to that in the primary analysis. For age at effect assessment, the estimated indirect effect increased until approximately 22 years of age and subsequently decreased. Uncertainty intervals were generally wider than those in the primary analysis.

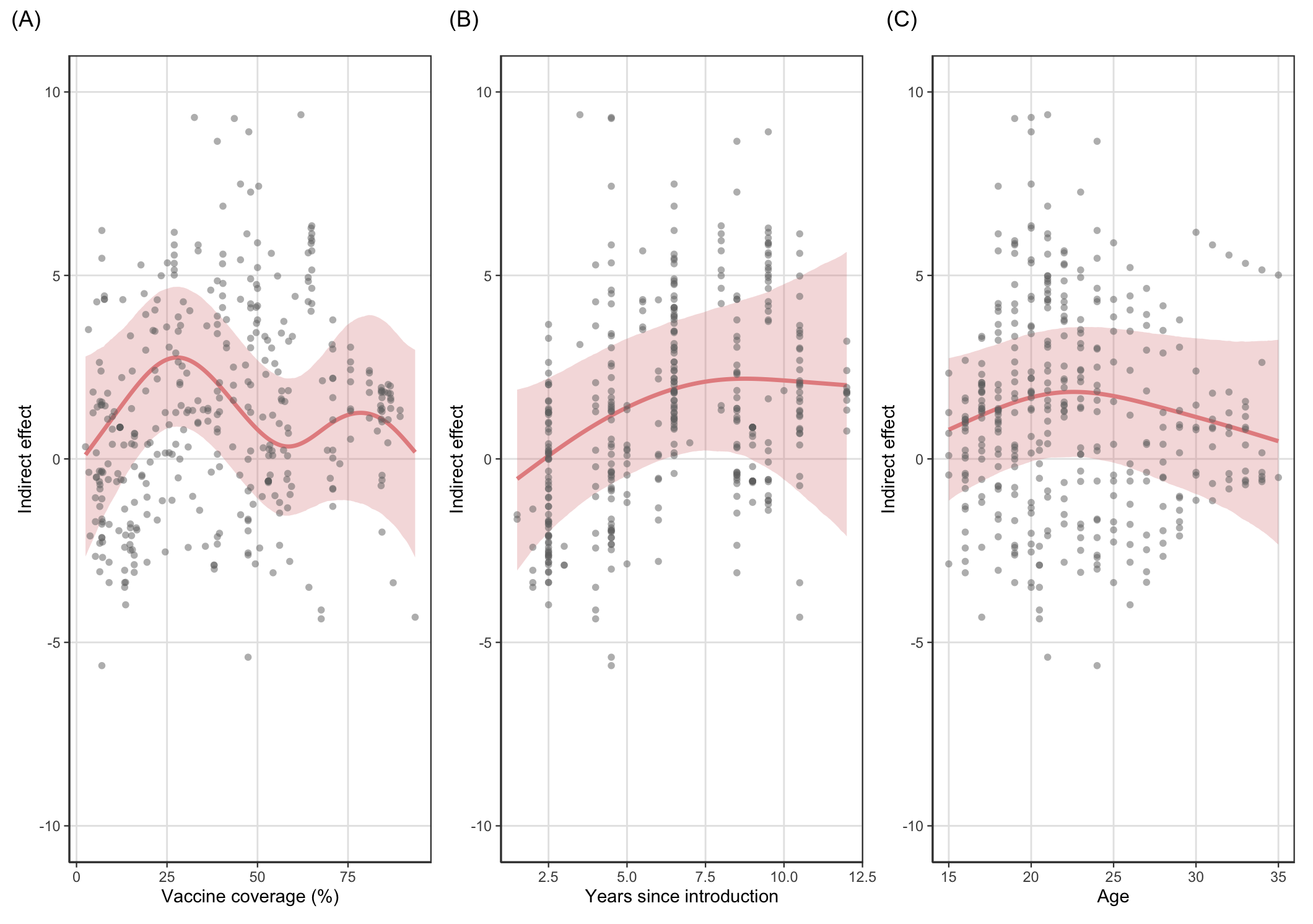

**Supplementary Figure 10. Predicted associations between the estimated indirect effect and (A) vaccine coverage (%), (B) time since vaccine introduction, and (C) age at effect assessment, after excluding observations from the US and UK.** Solid curves represent posterior mean estimates and shaded ribbons represent 95% credible intervals.

The relationships between years since vaccine introduction and the indirect effect across the three vaccination-eligibility groups showed similar patterns to those observed in the primary analysis (Supplementary Figure 11 B, E, and H). Some differences were observed for vaccine coverage and age at effect assessment. In the routine group, the association with vaccine coverage was more non-linear than in the primary analysis, with the estimated indirect effect initially increasing, declining after approximately 30% coverage, and showing greater variation at higher coverage levels (Supplementary Figure 11A). Age-related patterns also differed in the catch-up and non-targeted groups, with estimated indirect effects showing an inverted U-shaped pattern and reaching their highest values at approximately 30 years of age at effect assessment (Supplementary Figure 11F and I).

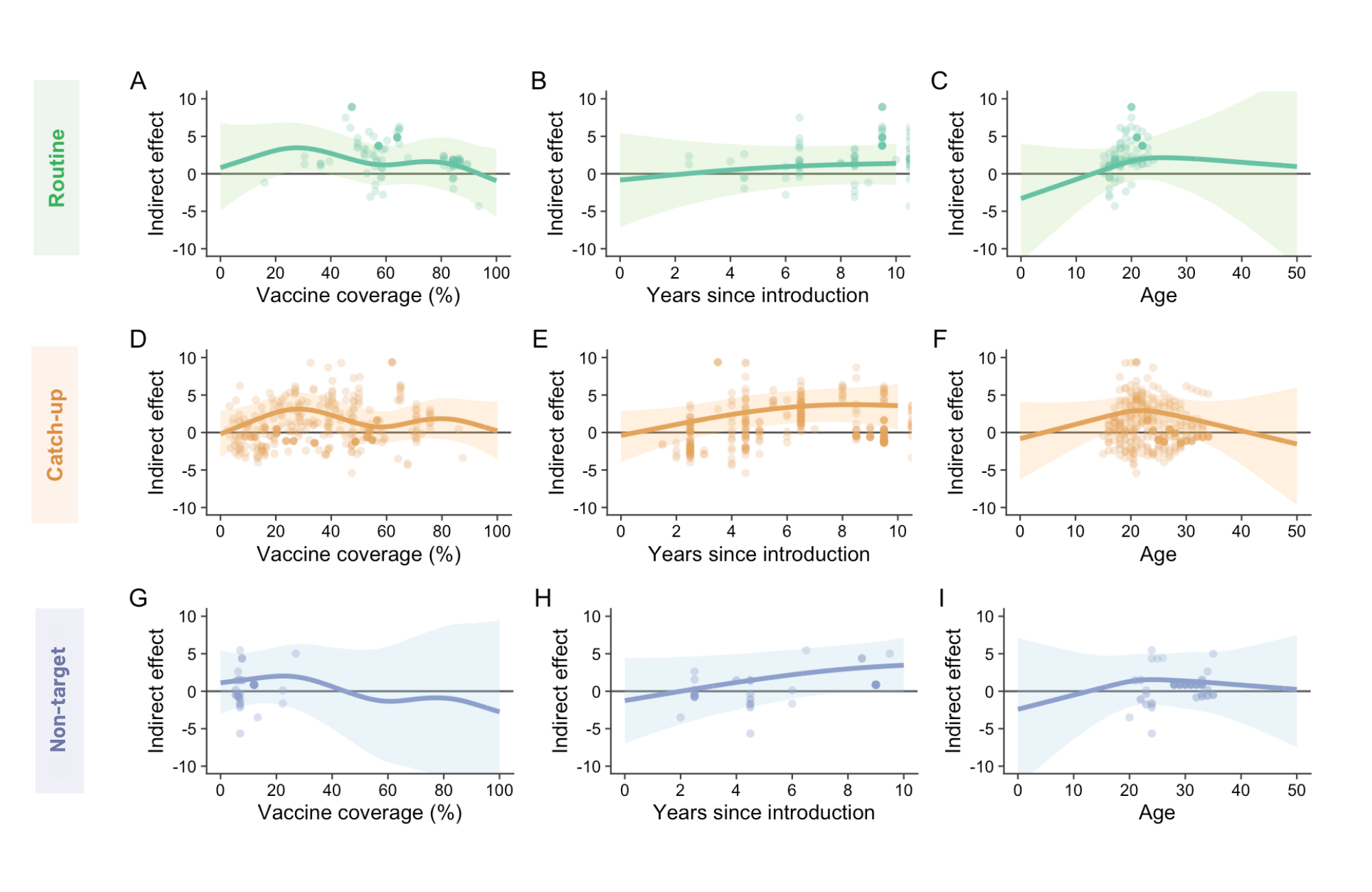

**Supplementary Figure 11. Predicted associations between the estimated indirect effect and vaccine coverage (%), time since vaccine introduction, and age at effect assessment across vaccination-eligibility groups after excluding observations from the US and UK.** (A–C) Routine group; (D–F) catch-up group; and (G–I) non-targeted group. Solid curves represent posterior mean estimates and shaded ribbons represent 95% credible intervals.

**6.2 Model Results US and UK Only**

When the analysis was restricted to observations from the US and UK, the overall associations were broadly consistent with those observed in the primary analysis (Supplementary Figure 12). However, steeper negative associations were observed for vaccine coverage and age at effect assessment, while the association with time since vaccine introduction remained similar.

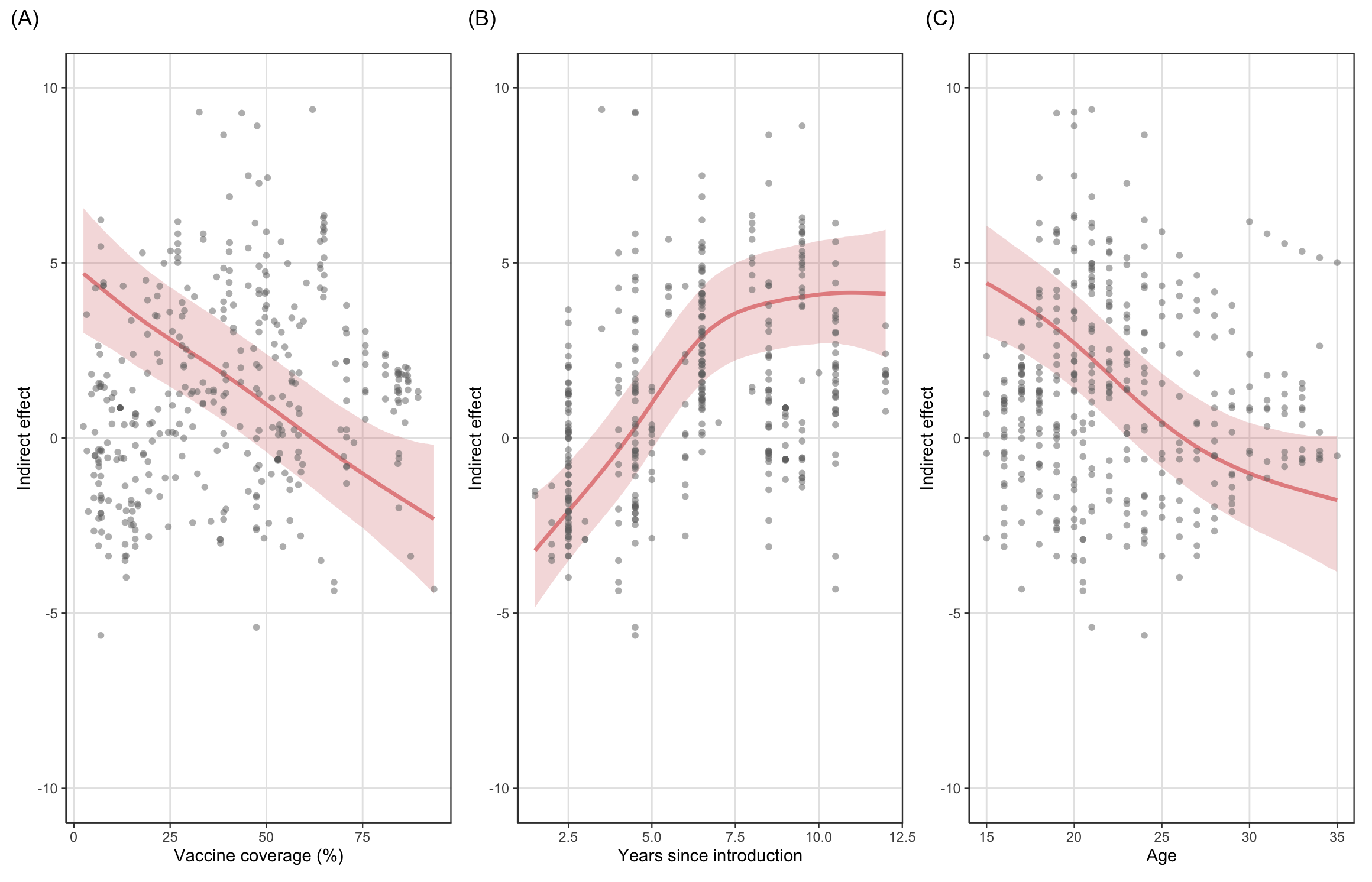

**Supplementary Figure 12. Predicted associations between the estimated indirect effect and (A) vaccine coverage (%), (B) time since vaccine introduction, and (C) age at effect assessment, using only observations from the US and UK.** Solid curves represent posterior mean estimates and shaded ribbons represent 95% credible intervals.

Across vaccination-eligibility groups, the associations with vaccine coverage and time since vaccine introduction were generally similar to those in the primary analysis (Supplementary Figure 13). Greater differences were observed for age at effect assessment. In the routine group, the estimated indirect effect showed a slight increase with age, whereas in the primary analysis it increased until approximately 20 years of age and then remained relatively stable. In the catch-up group, the estimated indirect effect declined more steeply with increasing age than in the primary analysis. In the non-targeted group, the positive association between age at effect assessment and the estimated indirect effect was broadly consistent with the primary analysis.

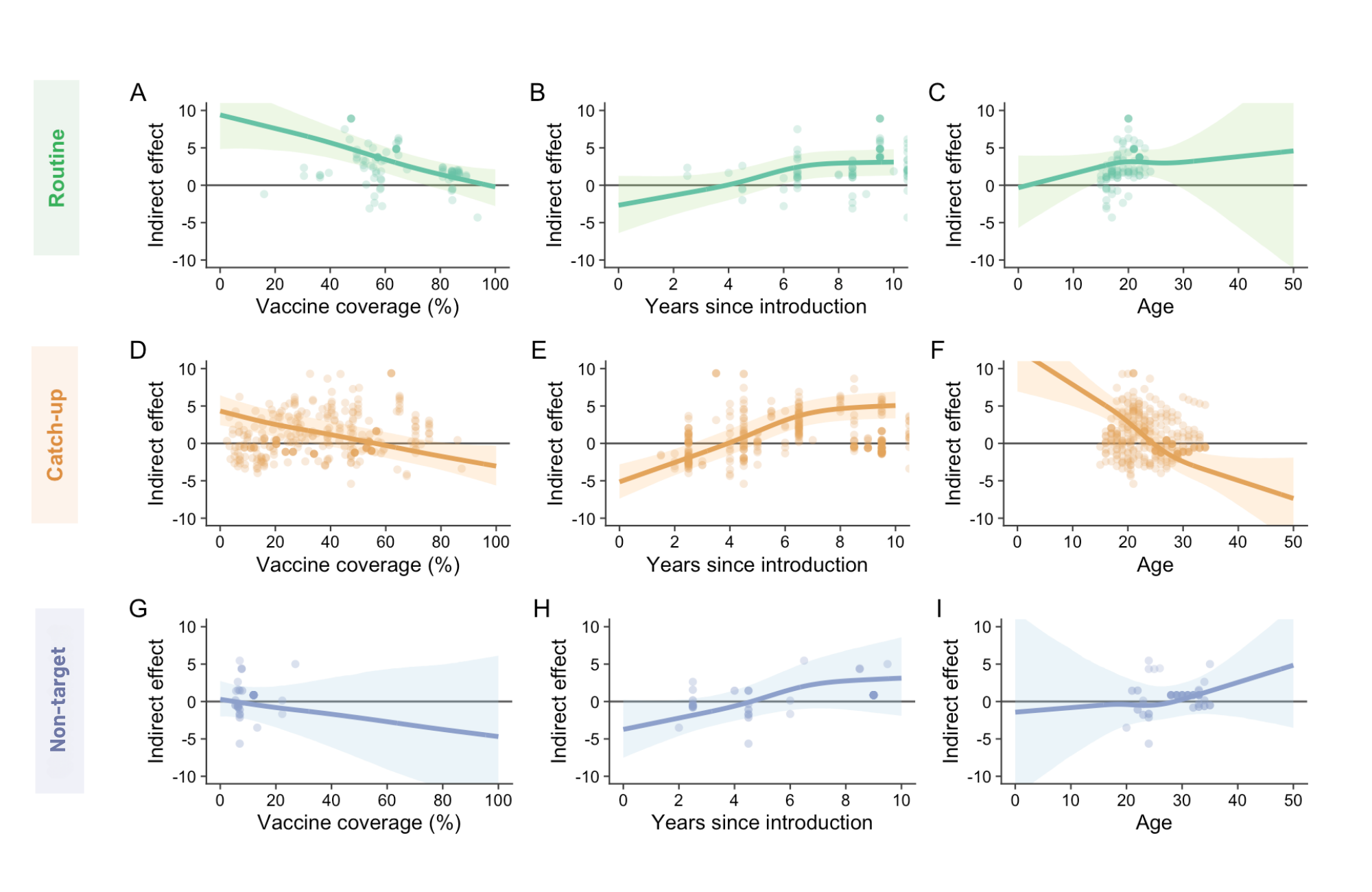

**Supplementary Figure 13. Predicted associations between the estimated indirect effect and vaccine coverage (%), time since vaccine introduction, and age at effect assessment across vaccination-eligibility groups using only observations from the US and UK.** (A–C) Routine group; (D–F) catch-up group; and (G–I) non-targeted group. Solid curves represent posterior mean estimates and shaded ribbons represent 95% credible intervals.

7. **Study-level random intercepts**

We plotted the study-level random intercepts to assess between-study heterogeneity after controlling for vaccine coverage, years since vaccine introduction, age, and vaccination-eligibility group. Most studies were centred around the overall population-level intercept. A few studies showed larger positive or negative deviations (Supplementary Figure 14).

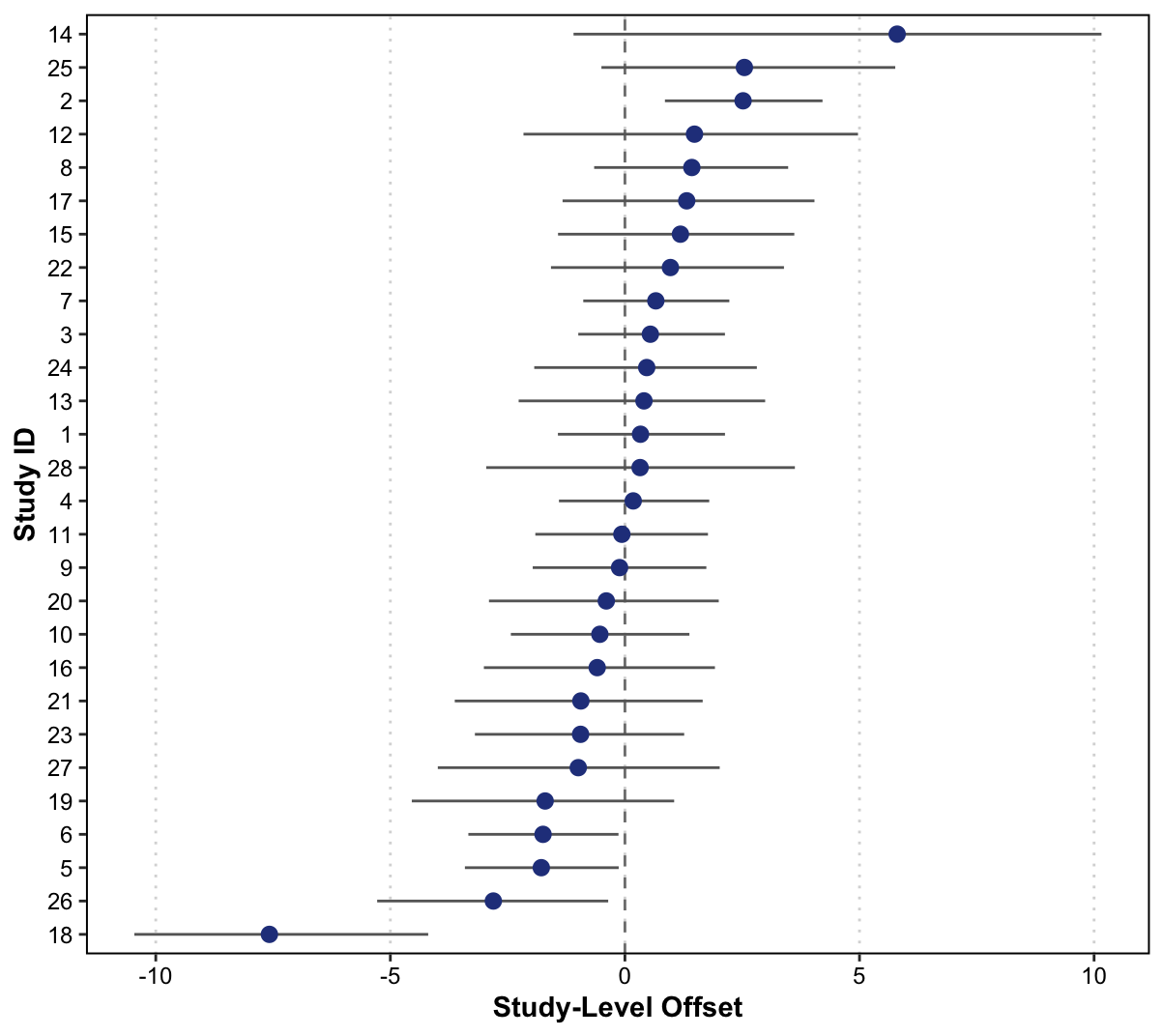

**Supplementary Figure 14. Study-level random intercepts**

8. **Vaccination schedule for each country/region**

We drew an HPV vaccination immunisation schedule for each country or region included in this study. The vaccination schedule was obtained from open-access literature, national vaccination guidelines, and government reports. The sources used for each country or region are listed in Supplementary Table 6. For each country-specific plot, colours were used to represent vaccination-eligibility group corresponding to each age group in each calendar year.

| **Country or region** | **Sources** |
| --- | --- |
| United Kingdom | ^6–8^ |
| United States | ^9^ |
| Australia | ^10–14^ |
| Denmark | ^15^ |
| Sweden | ^15–17^ |
| Norway | ^15^ |
| Japan | ^18,19^ |
| Netherlands | ^20^ |
| Finland | ^21^ |

**Supplementary Table 6. Sources used to reconstruct HPV vaccination programme schedules by country or region.**

**United Kingdom**

Catch-up for girls

Routine for girls

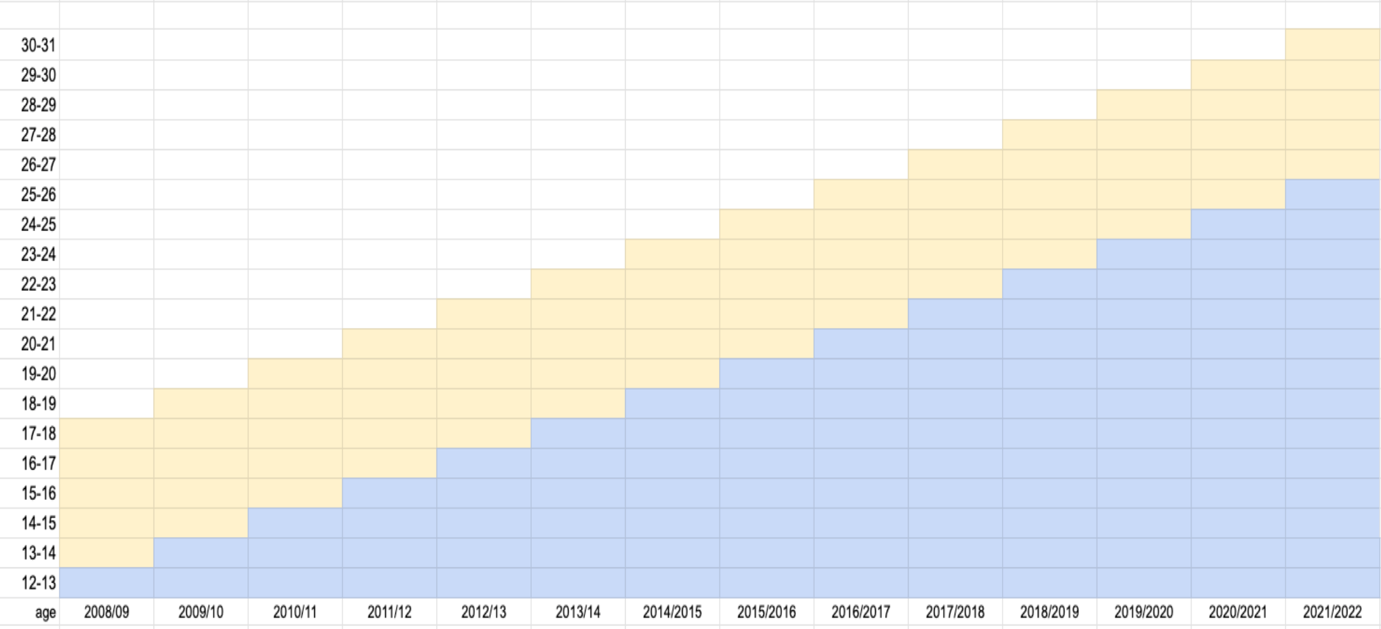

4-valent HPV vaccine

Routine: girls aged 12–13; catch-up: girls aged 13–18

2-dose schedule (<15 years)

2-valent HPV; 3 doses

**Supplementary Figure 15. HPV vaccination programme schedule in the United Kingdom.**

**United States**

Catch-up for girls

Routine for girls

Routine for boys

*

Catch-up for boys

\#

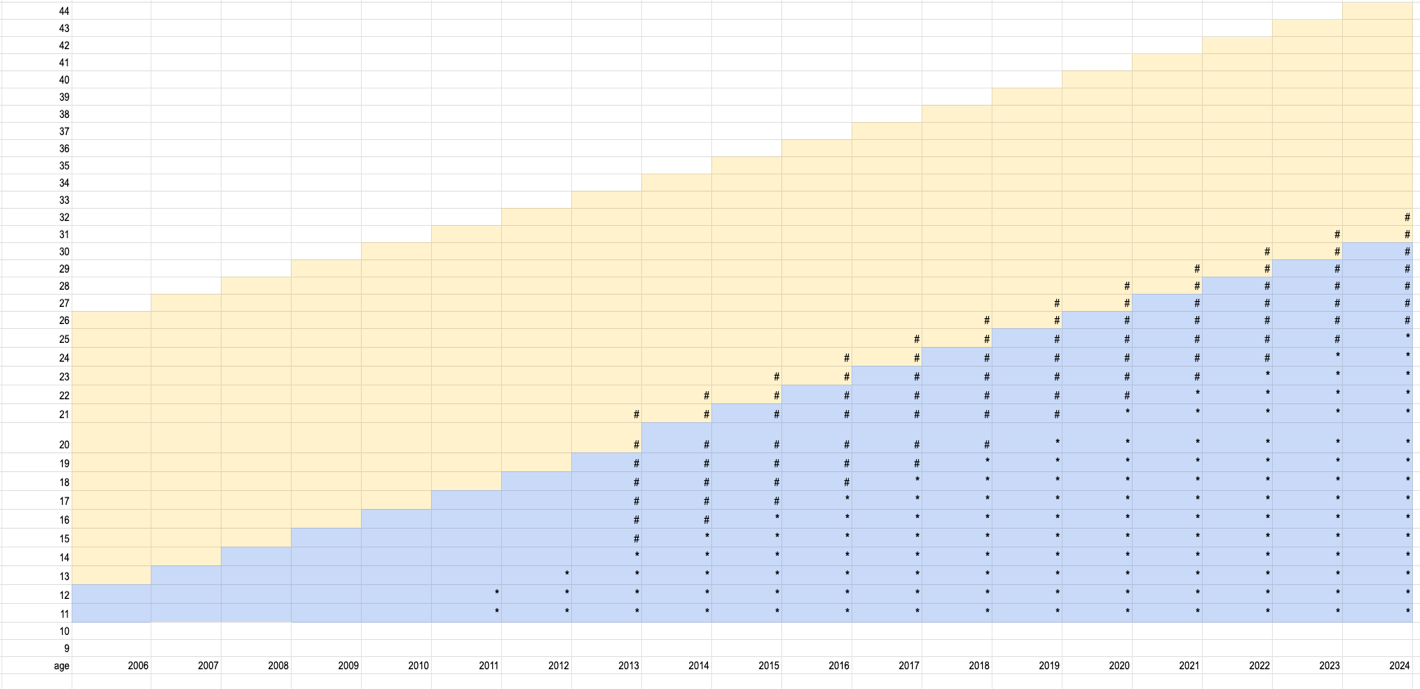

Routine and catch-up: boys

2-valent HPV vaccine

9-valent HPV vaccine

Routine: ages 11–12; catch-up: ages 13–26

4-valent HPV; 3 doses

**Supplementary Figure 16. HPV vaccination programme schedule in the United States.**

**Australia**

Catch-up for girls

Routine for girls

Routine for boys

*

Catch-up for boys

\#

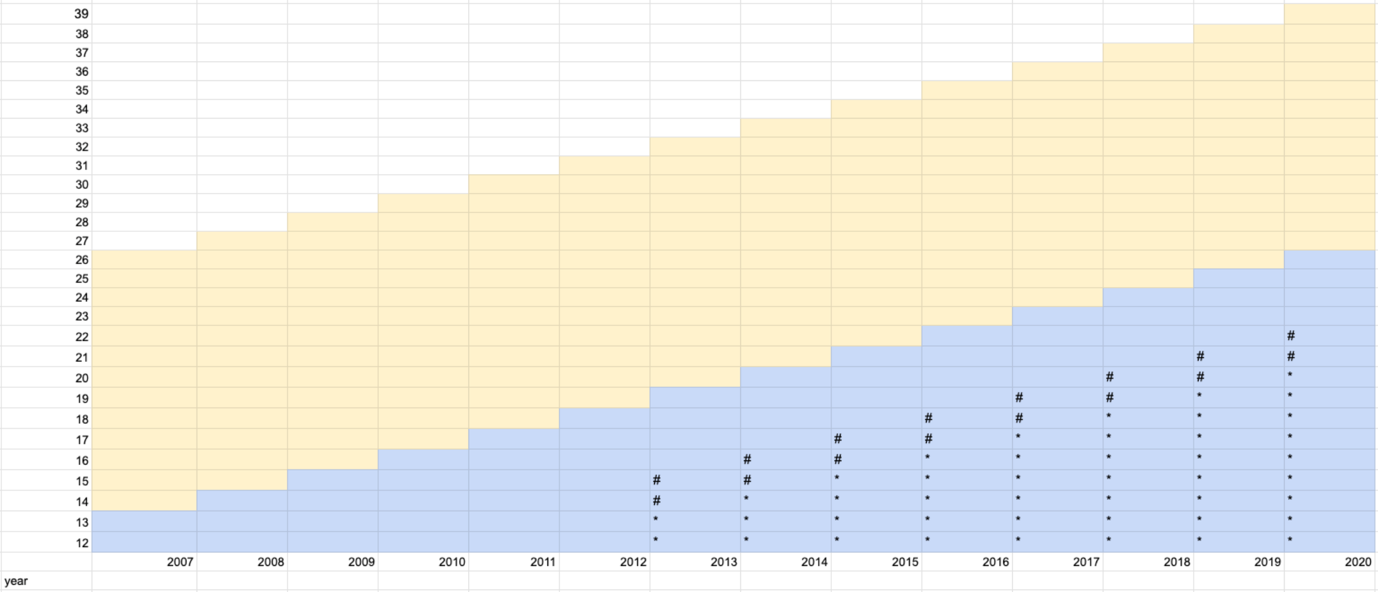

Catch-up: boys aged 14–15

Gender-neutral routine: ages 12–13

Routine: girls aged 12–13

Catch-up: girls aged 14–16

9-valent HPV; 2 doses

4-valent HPV; 3 doses

**Supplementary Figure 17. HPV vaccination programme schedule in Australia.**

**Denmark**

Catch-up for girls

Routine for girls

Second catch up

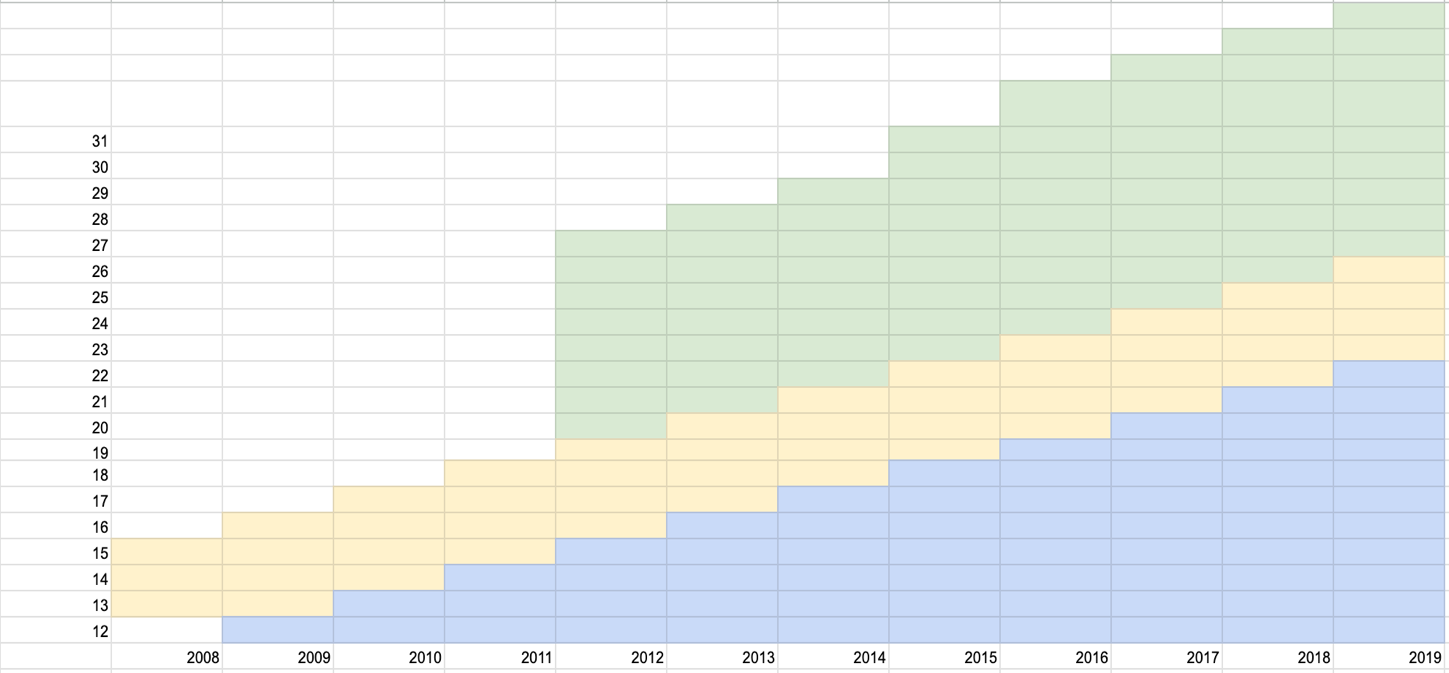

9-valent HPV vaccine

2-valent HPV vaccine

4-valent HPV vaccine

Second catch-up: women aged <27

Routine: girls aged 12

Catch-up: girls aged 13–15

**Supplementary Figure 18. HPV vaccination programme schedule in Denmark.**

**Sweden**

Catch-up for girls

Routine for girls

Subsidise

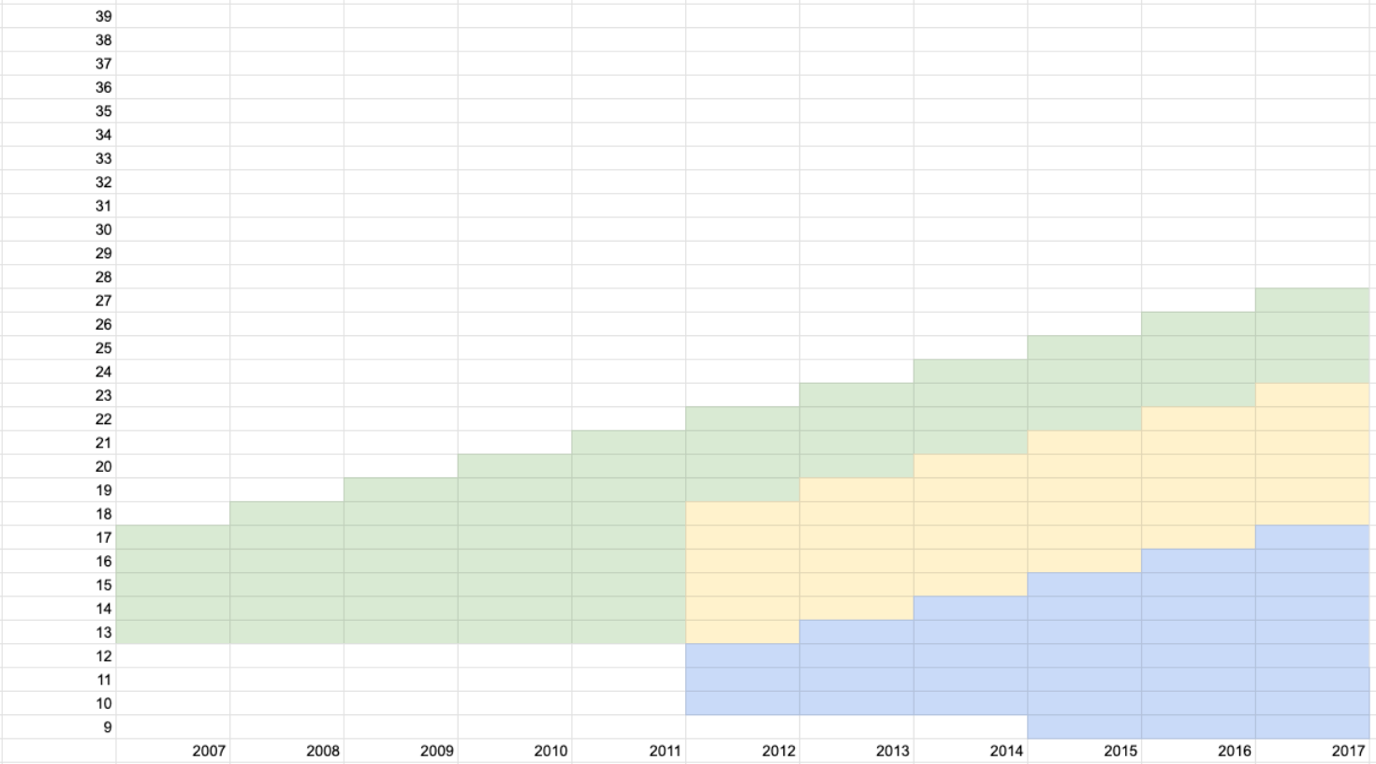

Subsidised vaccination: girls aged 13–17

School-based programme: girls aged 10–12; catch-up: girls aged 13–18

Two-dose schedule for girls aged 9–14

**Supplementary Figure 19. HPV vaccination programme schedule in Sweden.**

**Norway**

Catch-up for girls

Routine for girls

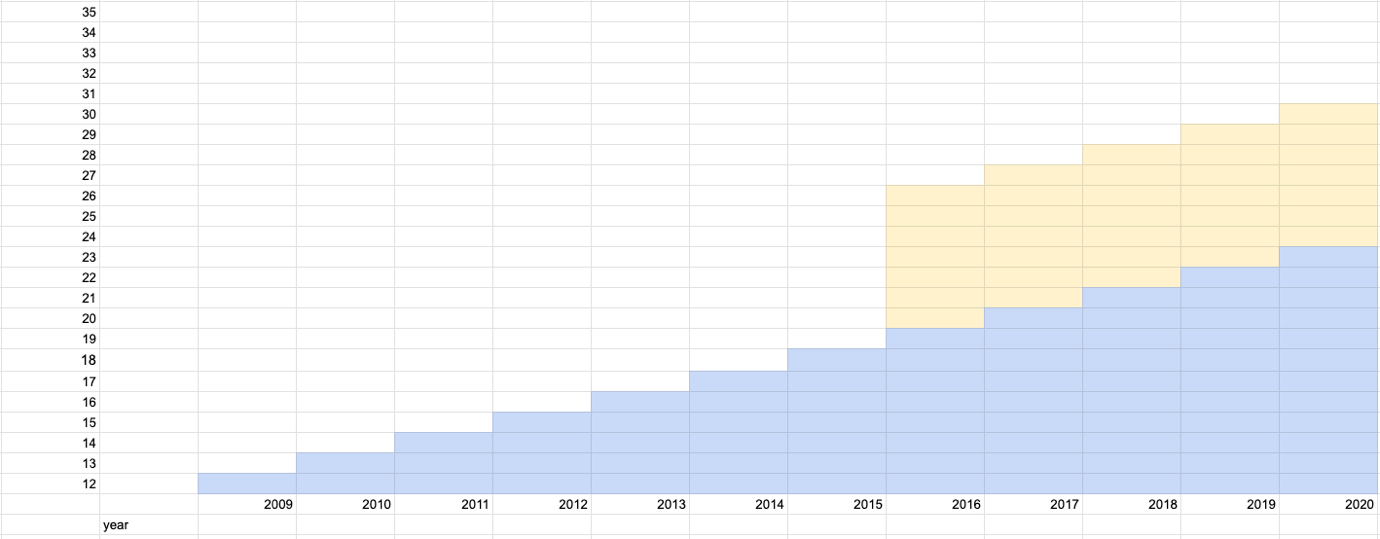

Routine: girls aged 12

Catch-up: girls and women up to age 26

**Supplementary Figure 20. HPV vaccination programme schedule in Norway.**

**Japan**

Catch-up for girls

Routine for girls

Subsidise

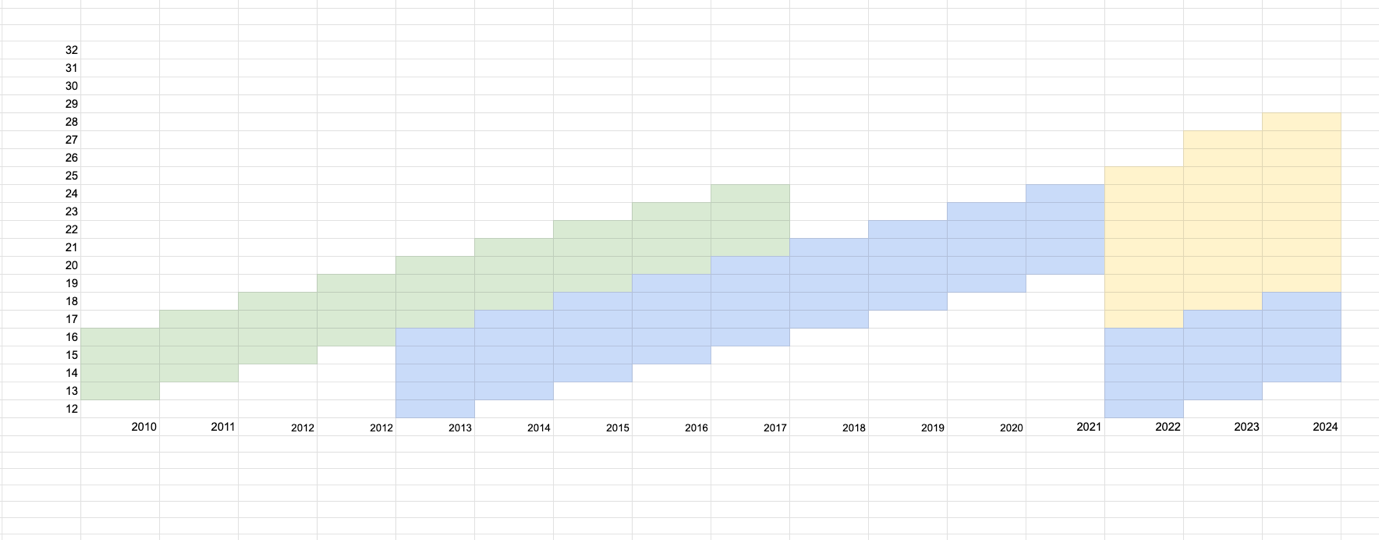

Routine: girls aged 12–16; catch-up: women aged 16–24

Routine: girls aged 12–16

**Supplementary Figure 21. HPV vaccination programme schedule in Japan.**

**Netherlands**

Catch-up for girls

Routine for girls

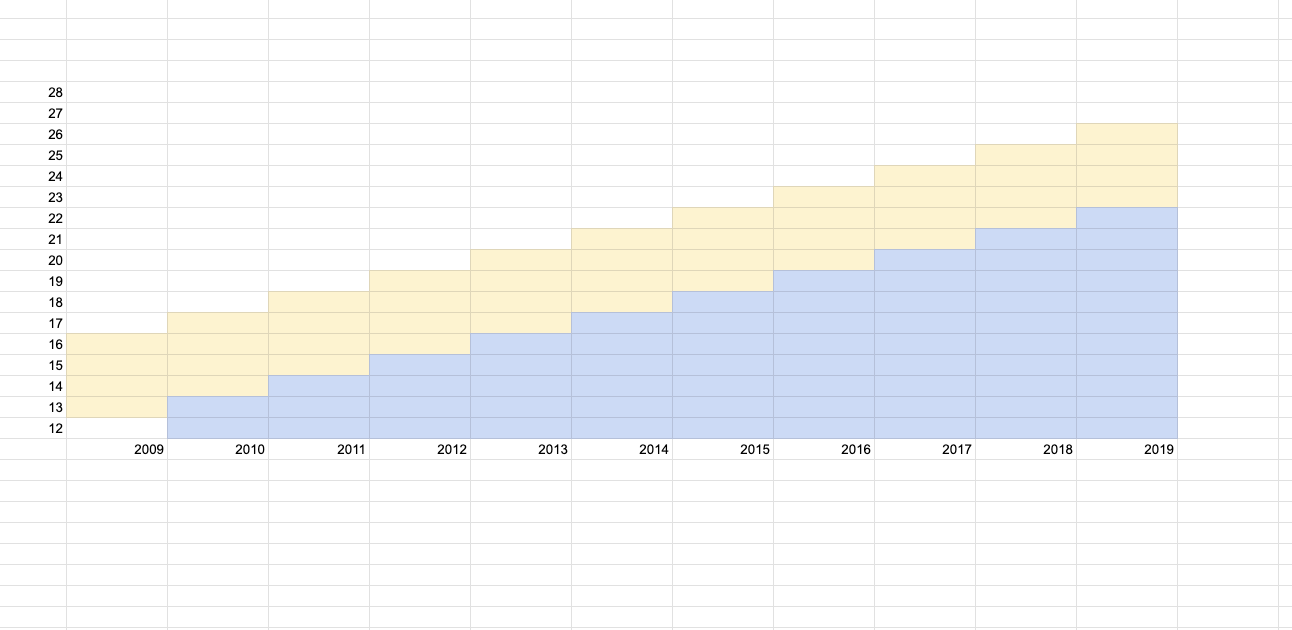

Routine: girls aged 12–13

Catch-up: girls aged 13–16

**Supplementary Figure 22. HPV vaccination programme schedule in the Netherlands.**

**Finland**

Catch-up for girls

Routine for girls

Routine for boys

*

Catch-up for boys

\#

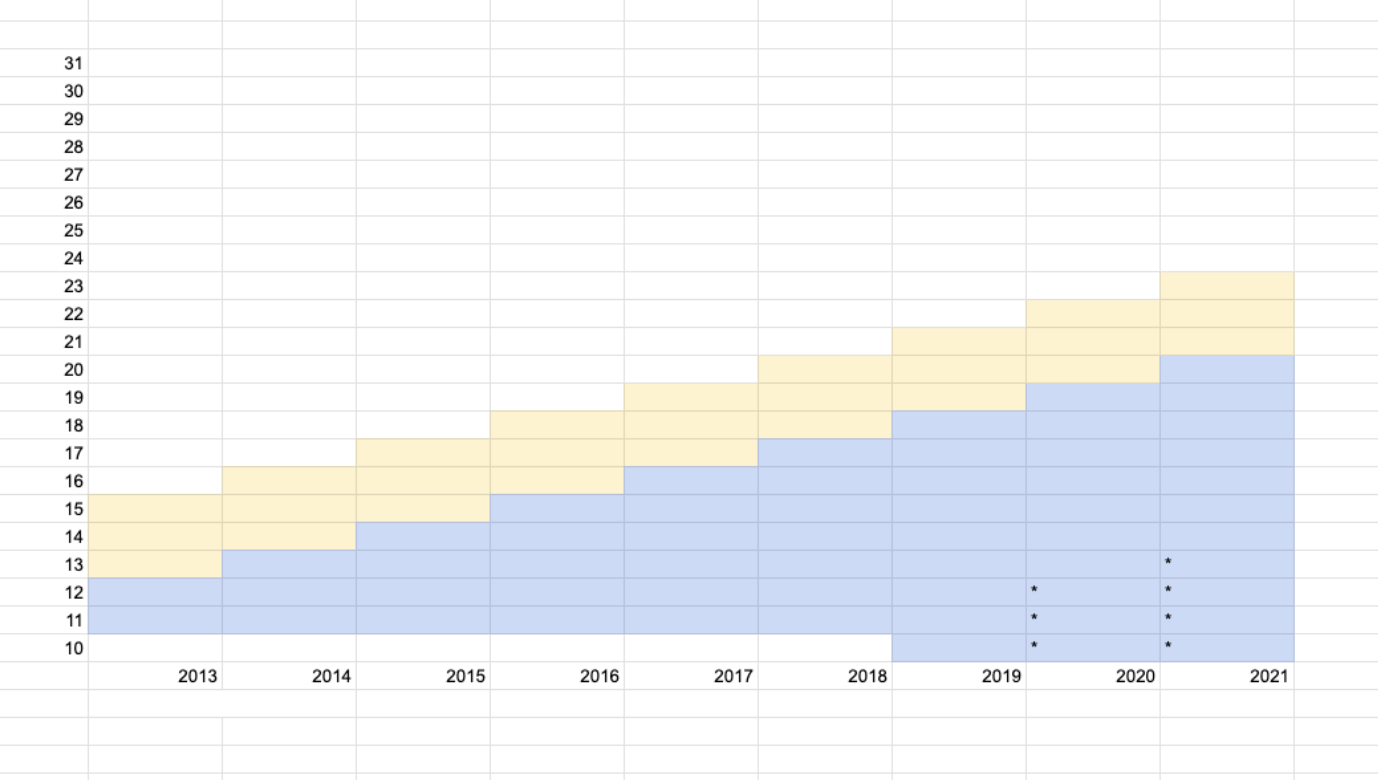

Routine: girls aged 11–12

Routine: girls aged 10–12

Routine: boys aged 10–12

Catch-up: girls aged 13–15

**Supplementary Figure 23. HPV vaccination programme schedule in Finland.**
